# Dense phenotyping from electronic health records enables machine-learning-based prediction of preterm birth

**DOI:** 10.1101/2020.07.15.20154864

**Authors:** Abin Abraham, Brian Le, Idit Kosti, Peter Straub, Digna R. Velez-Edwards, Lea K. Davis, J. M. Newton, Louis J. Muglia, Antonis Rokas, Cosmin A. Bejan, Marina Sirota, John A. Capra

## Abstract

Identifying pregnancies at risk for preterm birth, one of the leading causes of worldwide infant mortality, has the potential to improve prenatal care. However, we lack broadly applicable methods to accurately predict preterm birth risk. The dense longitudinal information present in electronic health records (EHRs) is enabling scalable and cost-efficient risk modeling of many diseases, but EHR resources have been largely untapped in the study of pregnancy. Here, we apply machine learning to diverse data from EHRs to predict singleton preterm birth. Leveraging a large cohort of 35,282 deliveries, we find that machine learning models based on billing codes alone can predict preterm birth risk at various gestational ages (e.g., ROC-AUC=0.75, PR-AUC=0.40 at 28 weeks of gestation) and outperform comparable models trained using known risk factors (e.g., ROC-AUC=0.65, PR-AUC=0.25 at 28 weeks). Examining the patterns learned by the model reveals it stratifies deliveries into interpretable groups, including high-risk preterm birth sub-types enriched for distinct comorbidities. Our machine learning approach also predicts preterm birth sub-types (spontaneous vs. indicated), mode of delivery, and recurrent preterm birth. Finally, we demonstrate the portability of our approach by showing that the prediction models maintain their accuracy on a large, independent cohort (5,978 deliveries) from a different healthcare system. By leveraging rich phenotypic and genetic features derived from EHRs, we suggest that machine learning algorithms have great potential to improve medical care during pregnancy.

## Introduction

Preterm birth, occurring before 37 weeks of completed gestation, affects approximately 10% of pregnancies globally[1–3] and is the leading cause of infant mortality worldwide[4,5]. The causes of preterm birth are multifactorial, since different biological pathways and environmental exposures can trigger premature labor[6]. Large epidemiological studies have identified many risk factors, including multiple gestations[1], cervical anatomic abnormalities[7], and maternal age[8]. Notably, even though a history of preterm birth [9] is one of the strongest risk factors, the recurrence rate remains low at < 30%[10,11]. Additionally, maternal race is associated with risk for preterm birth; Black women have twice the prevalence compared to white women[1,12]. Preterm births have a heterogenous clinical presentation and cluster based on maternal, fetal, or placental conditions[3]. These obstetric and systemic comorbidities (e.g. pre-existing diabetes, cardiovascular disease) can also increase risk for preterm birth[13,14].

Despite our understanding of numerous risk factors, there are no accurate methods to predict preterm birth. Some biomarkers associate with preterm birth, but their best performance is limited to a subset of all cases[15,16]. Recently, analysis of maternal cell-free RNA and integrated -omic models have emerged as promising approaches[17–19], but initial results were based on a small pregnancy cohort and require further validation. In silico classifiers based on demographic and clinical risk factors have the advantage of not requiring serology or invasive testing. However, even in large cohorts (>1 million individuals), demograpic- and risk-factor-based models report limited discrimination (AUC=0.63-0.74)[20–23]. To date, we lack effective screening tools and preventative strategies for prematurity[24].

Electronic health records (EHRs) are scalable, readily available, and cost-efficient for disease-risk modeling[25]. EHRs capture longitudinal data across a broad set of phenotypes with detailed temporal resolution. EHR data can be combined with socio-demographic factors and family medical history to comprehensively model disease risk[26–28]. EHRs are also increasingly being augmented by linking patient records to molecular data, such as DNA and laboratory test results[29]. Since preterm birth has a substantial heritable risk[30], combining rich phenotypes with genetic risk may lead to better prediction.

Machine learning models have shown promise for accurate risk stratification across a variety of clinical domains[31–33]. However, despite the rapid adoption of machine learning in translational research, a review of 107 risk prediction studies reported that most models used only few variables, did not consider longitudinal data, and rarely evaluated model performance across multiple sites[34]. Studies using machine learning to predict preterm birth have relied on small cohorts, subsets of preterm birth, and are rarely replicated in external datasets[22,35–37]. Pregnancy research is especially well poised to benefit from machine learning approaches[26]. Per standard of care during pregnancy, women are carefully monitored with frequent prenatal visits, medical imaging, and clinical laboratories tests. Compared to other clinical contexts, pregnancy and the corresponding clinical surveillance occur in a defined timeframe based on gestational length. Thus, EHRs are well-suited for modeling pregnancy complications, especially when combined with the well documented outcomes at the end of pregnancy.

In this study, we combine multiple sources of data from EHRs to predict preterm birth using machine learning. From Vanderbilt’s EHR database (>3.2 million records) and linked genetic biobank (>100,000 individuals), we identified a large cohort of women (n=35,282) with documented deliveries. We trained models (gradient boosted decision trees) that combine demographic factors, clinical history, laboratory tests, and genetic risk with billing codes (ICD-9 and CPT) to predict preterm birth. We find models trained on only billing codes show potential for predicting preterm birth. Billing code based models outperform a similar model using only known clinical risk factors. Across a variety of clinical contexts, such as second or spontaneous preterm birth, our models maintain accuracy. By investigating the patterns learned by our models, we identify clusters with distinct preterm birth risk and comorbidity profiles.

Finally, we demonstrate the generalizability of our billing-code-based models on an external, independent cohort from the University of California, San Francisco (UCSF, n=5,978). Prediction models trained at Vanderbilt maintain high accuracy in the external cohort with only a modest drop in performance. Our findings provide a proof-of-concept that machine learning on rich phenotypes in EHRs show promise for portable, accurate, and non-invasive prediction of preterm birth. The strong predictive performance across clinical context and preterm birth subtypes argues that machine learning models have the potential to add value during the management of pregnancy; however, further work is needed before these models can be applied in clinical settings.

## Results

### Assembling pregnancy cohort and ascertaining delivery type from Vanderbilt EHRs

From the Vanderbilt EHR database (>3.2 million patients), we identified a ‘delivery cohort’ of 35,282 women with at least one delivery in the Vanderbilt hospital system (Fig. 1A). In addition to ICD and CPT billing codes, we extracted demographic data, past medical histories, obstetric notes, clinical labs, and genome-wide genetic data for the delivery cohort. Because billing codes were the most prevalent data in this cohort (n=35,282), we quantified the pairwise overlap between billing codes and each other data type. The largest subset included women with billing codes paired with demographic data (n=33,570). The smallest subset was women with billing codes paired with genetic data (n=905; Fig. 1C). The mean maternal age at the first delivery in the delivery cohort was 27.3 years (23.0–31.0 years, 25^th^ and 75^th^ percentiles, Fig. S1A). The majority of women in the cohort self- or third-party reported as white (n=21,343), Black (n=6,178), or Hispanic (n=3,979; Fig. S1B). The estimated gestational age (EGA) distribution had a mean of 38.5 weeks (38.0 to 40.3 weeks, 25th to 75th percentile; Fig. 1D). The rate of multiple gestations (e.g. twins, triplets) was (7.6%, n=1,353). Since multiple gestation pregnancies are more likely to deliver preterm, we developed prediction models using singleton pregnancies unless otherwise stated.

**Figure 1.**
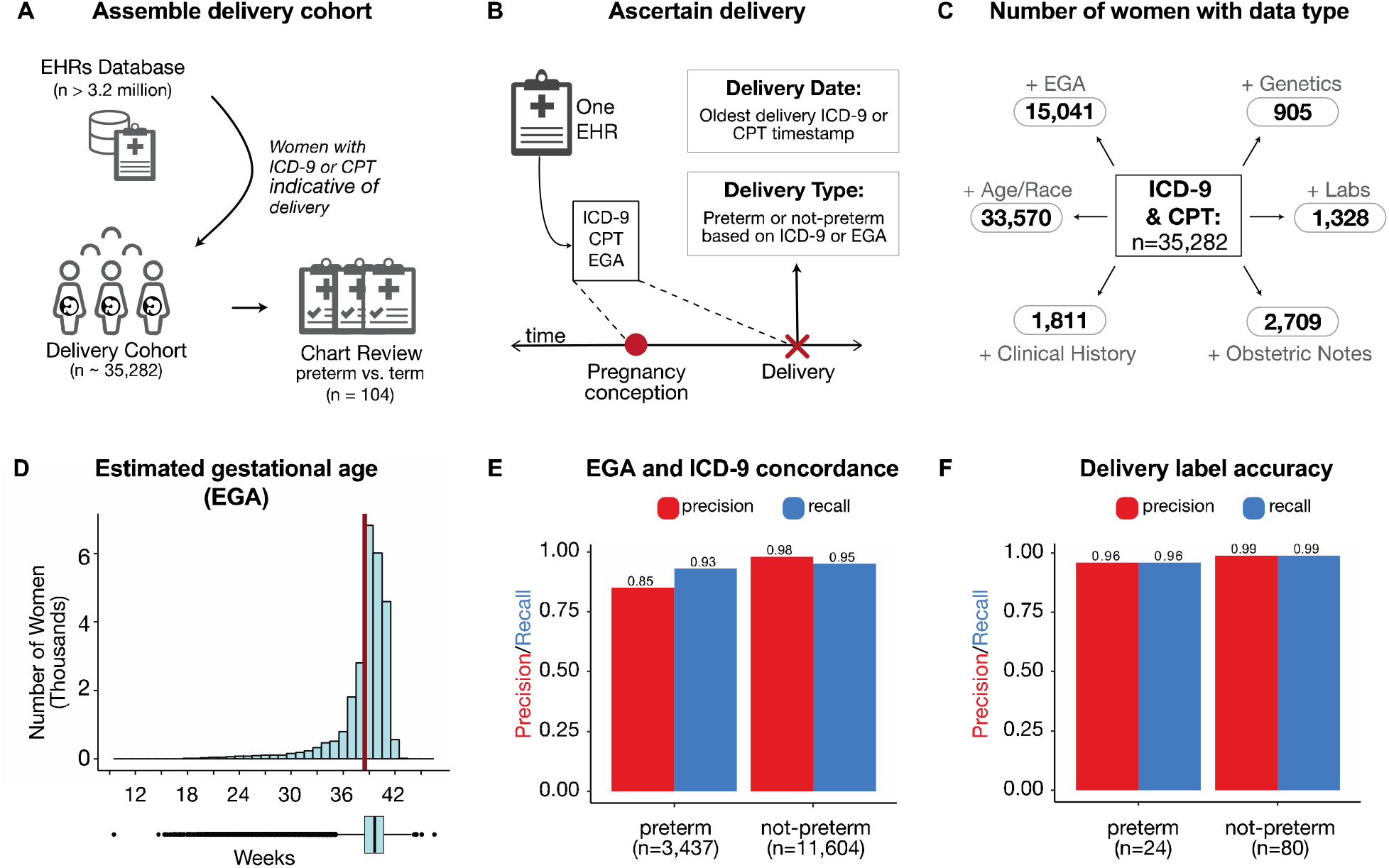
Definition and attributes of Vanderbilt delivery cohort. **(A)** Schematic overview of the assembly of the delivery cohort from electronic health records (EHRs). Using billing codes, women with at least one delivery were extracted from the EHR database (n=35,282). **(B)** Delivery date and type were ascertained using ICD-9, CPT, and/or estimated gestational age (EGA) from each woman’s EHR (Methods). From this cohort, 104 randomly selected EHRs were chart reviewed to validate the preterm birth label for the first recorded delivery. **(C)** Number of women in billing code cohort with estimated gestational age (+EGA), demographics (+Age, self- or third-party reported Race), clinical labs (+Labs), clinical obstetric notes (+Obstetric notes), patient clinical history (+Clinical History), and genetic data (+Genetics). **(D)** The EGA distribution at delivery (mean 38.5 weeks (red line); 38.0-40.3 weeks, 25^th^ and 75^th^ percentiles). Less than 0.015% (n=49) deliveries have EGA below 20 weeks. **(E)** The concordance between estimated gestational age (EGA) within three days of delivery and ICD-9 based delivery type for the 15,041 women with sufficient data for both. Precision and recall values were > 93% across labels except for preterm precision (85%). **(F)** Accuracy of delivery type phenotyping. The phenotyping algorithm was evaluated by chart review of 104 randomly selected women. The approach has high precision and recall for binary classification of ‘preterm’ or ‘not-preterm’.

To determine the delivery date and type (preterm vs. not-preterm) at scale across our large cohort, we developed a phenotyping algorithm using delivery-specific billing codes and estimated gestational age at delivery. For women with multiple pregnancies, we only considered the earliest delivery. We find that delivery-specific billing codes that can be used to label preterm births have high concordance (PPV≥0.85, Recall ≥0.95) with EGA based delivery labels (Fig. 1E). Our final algorithm combined billing codes and EGA when available (n=15,041, Fig. 1C). To evaluate the accuracy of the ascertained delivery labels, a domain expert blinded to the delivery type reviewed clinical notes from 104 EHRs selected at random from the delivery cohort. The algorithm had high accuracy: precision (positive predictive value) of 96% and recall (sensitivity) of 96% using the chart reviewed label as the gold standard (Fig. 1F).

### Boosted decision trees using billing codes identify preterm deliveries

Using this richly phenotyped delivery cohort, we evaluated how well the entire clinical phenome, defined as billing codes (ICD-9 and CPT) before and after delivery, could identify preterm births. With counts of each billing code (excluding those used to ascertain delivery type), we trained gradient boosted decision trees[38] to classify each mother’s first delivery as preterm or not-preterm. Boosted decisions trees are well-suited for EHR data because they require minimal transformation of the raw data, are robust to correlated features, and capture non-linear relationships[39]. Moreover, boosted decision trees have been successfully applied on a variety of clinical tasks[27,40,41].

In all evaluations, we held out 20% of the cohort as a test set and used the remaining 80% for training and validation (Fig. 2A). Boosted decision tree models trained on ICD-9 and CPT codes accurately identified preterm births (singletons and multiple gestations) with PR-AUC=0.86 (chance=0.22) and ROC-AUC=0.95 (Fig. S2A, B). While the combined ICD-9 and CPT based model achieved the best performance, models trained on either ICD-9 or CPT individually also performed well (PR-AUC ≥0.82; chance=0.22, ROC-AUC ≥0.93). All three models demonstrated good calibration with low Brier scores (≤0.092; Fig. S2C). Thus, billing codes across an EHR show potential as a discriminatory feature for *predicting* preterm birth.

**Figure 2.**
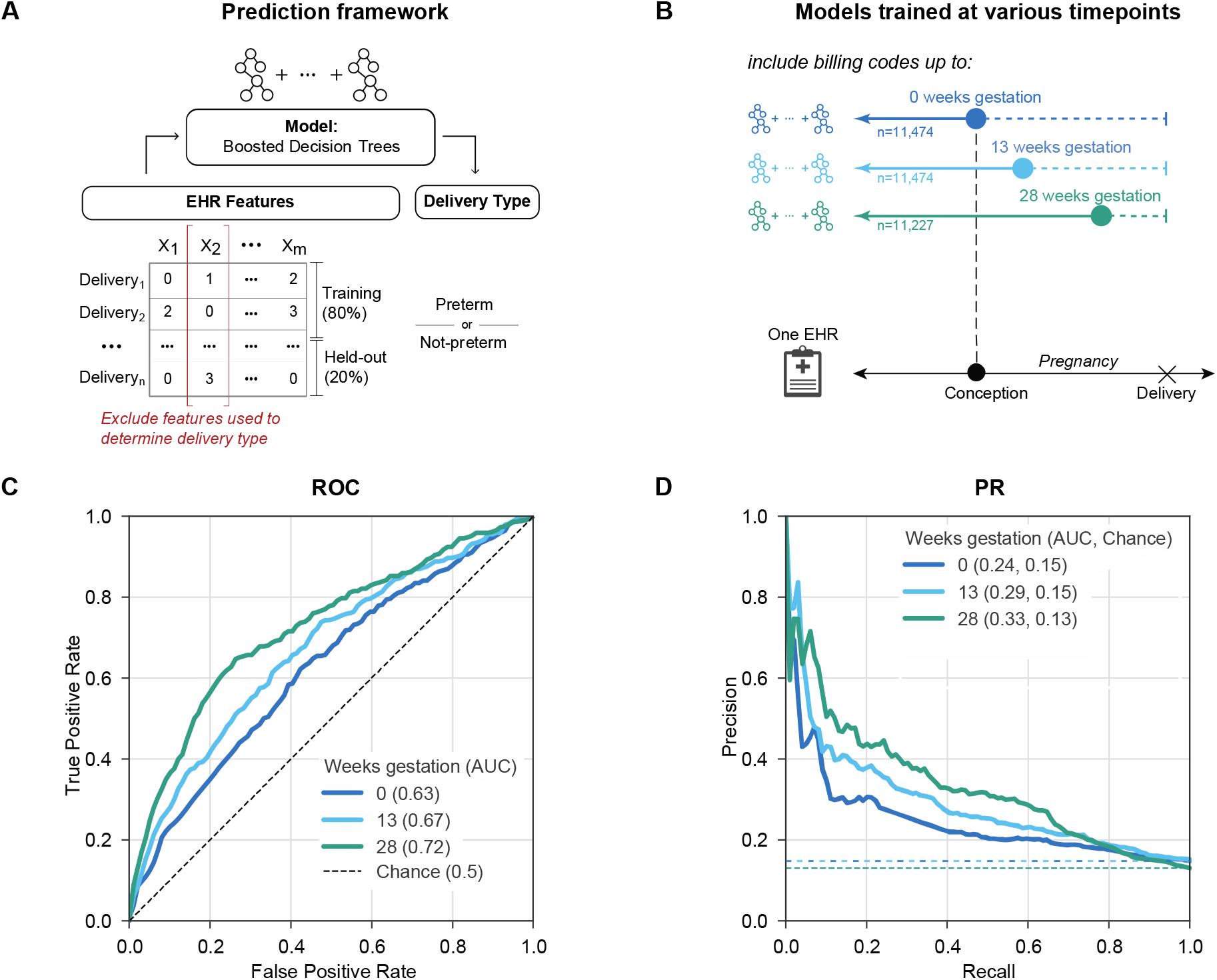
Machine learning classifiers accurately predict preterm birth using billing codes present before 28 weeks of gestation. **(A)** Machine learning framework for training and evaluating all models. We train models (boosted decision trees) on 80% of each cohort to predict the delivery as preterm or not-preterm. EHR features used to ascertain delivery type are excluded from training. Performance is reported on the held-out cohort consisting of 20% of deliveries using area under the ROC and precision-recall curves (ROC-AUC, PR-AUC). **(B)** We trained models using billing codes (ICD-9 and CPT) present before each of the following timepoints during pregnancy: 0, 13, and 28 weeks of gestation. These timepoints were selected to approximate gestational trimesters. Women who already delivered were excluded at each timepoint. To facilitate comparison across timepoints, we downsampled cohorts available so that the models were trained on a cohort with similar numbers of women (n=11,227 to 11,474). **(C)** The ROC-AUC increased from conception at 0 weeks (0.63, dark blue line) to 28 weeks of gestation (0.72, green line) compared to a chance (black dashed line) AUC of 0.5. **(D)** The model at 28 weeks of gestation achieved the highest PR-AUC (0.33). This is an underestimate of the possible performance; the accuracy improves further when all women with data available at 28 weeks are considered (Fig. 4B,C). Chance (dashed lines) represents the preterm birth prevalence in each cohort.

### Accurate prediction of preterm birth at 28 weeks of gestation

To evaluate preterm birth prediction in a clinical context, we trained a boosted decision tree model (Fig. 2A) on billing codes present before each of the following timepoints: 0, 13, and 28 weeks of gestatio (Fig. 2B). These timepoints were selected to approximately reflect pregnancy trimesters. We downsampled to achieve comparable number of singleton deliveries across each timepoint (n=11,227 to 11,474) to mitigate sample size as a potential confounder while comparing performance. We only considered active pregnancies at each timepoint; for example, a delivery at 27 weeks would not be included in the 28 week model, since the outcome would already be known. The ROC-AUC increased from conception (0 weeks; 0.63) to the highest performance at 28 weeks (0.72; Fig. 2C). The PR-AUC (Fig. 2D), which accounts for preterm birth prevalence, is highest at 28 weeks (0.33, chance=0.13). However, as we show in the next section, this is an underestimate of the ability to predict preterm delivery at 28 weeks due to the down-sampling of the number of training examples. As expected, when we included multiple gestations, the model performed even better (PR-AUC=0.42 at 28 weeks, chance=0.14; Fig. S3). Results were similar when models were trained using billing codes available before different timepoints from the date of delivery (Fig. S4).

To test whether differences in contact with the health system between cases and controls were driving performance, we trained a classifier based on the total number of codes in an individual’s EHR before delivery to predict preterm birth. This simple classifier failed to discriminate between delivery types with PR-AUC and ROC-AUC only slightly higher than chance (PR-AUC=0.19, chance=0.19; ROC-AUC=0.56, chance=0.5, Fig. S5). Therefore, cumulative disease burden or the number of contacts alone are not informative for predicting preterm birth.

### Integrating other EHR features does not improve model performance

In addition to billing codes, EHRs capture aspects of an individual’s health through different types of structured and unstructured data. We tested whether incorporating additional features from EHRs can improve preterm birth prediction. Models were evaluated using data available at 28 weeks of gestation; we selected this time point as a tradeoff between being sufficiently early for some potential interventions and late enough for sufficient data to be present to enable accurate predictions using billing codes. From the EHRs, we extracted sets of features including demographic variables (age, race), clinical keywords from obstetric notes, clinical lab tests ran during the pregnancy, and predicted genetic risk (polygenic risk score for preterm birth). To measure the performance gain for each feature set, we compared models trained using: the feature set only, billing codes only, and billing codes combined with the feature set (Fig. 3A). Within each feature set, the same pregnancies comprised the training and held-out sets for the three models. However, the number of deliveries (training + held-out sets) varied widely across feature sets (n=462 to 20,342) due to the differing availability of each feature type.

**Figure 3.**
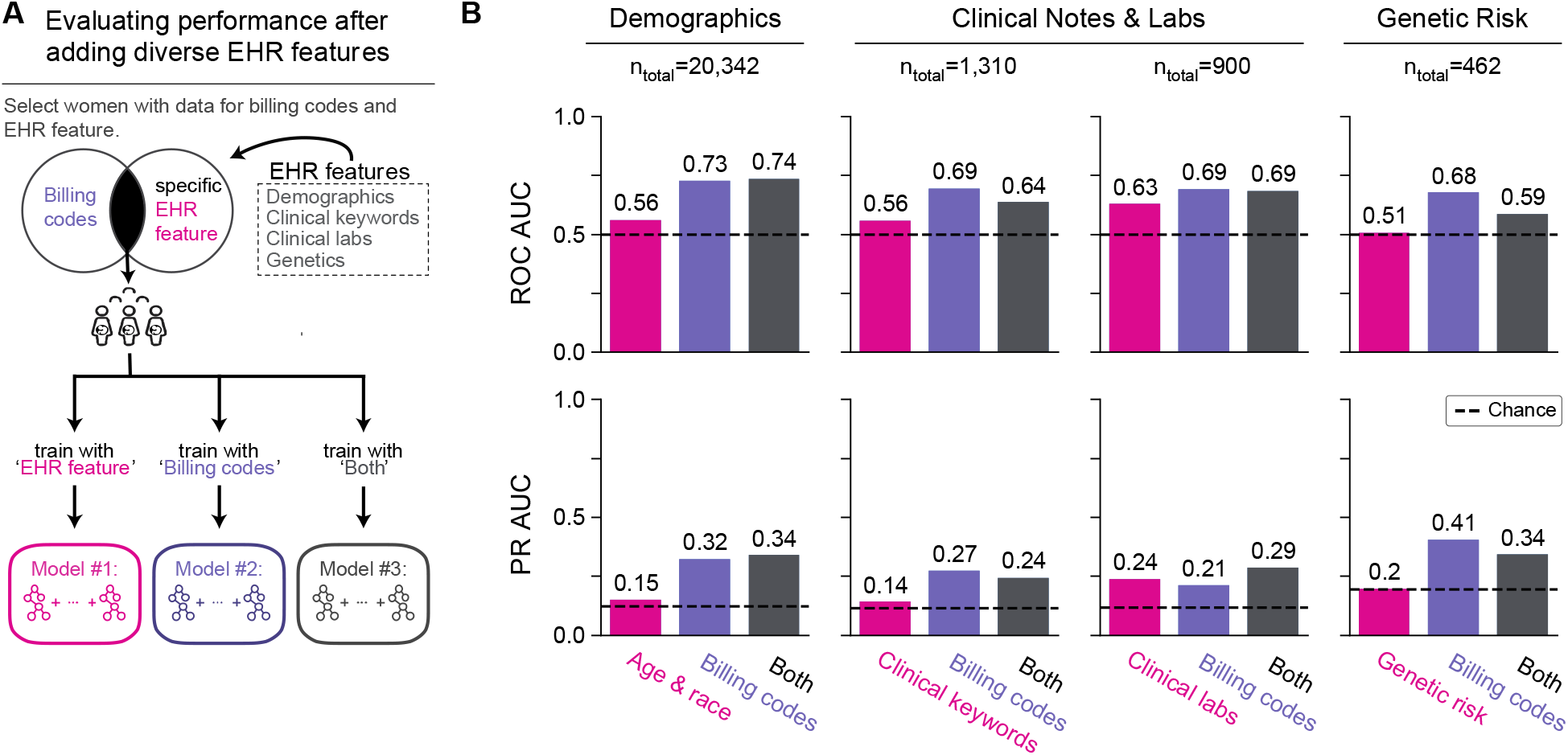
Combing demographic, clinical, and genetic features does not substantially improve preterm birth prediction compared to using only billing codes. **(A)** Framework for evaluating change in preterm birth prediction performance after incorporating diverse types of EHR features with billing codes (ICD-9 and CPT codes). We used only features and billing codes occurring before 28 weeks of gestation. EHR features are grouped by sets of demographic factors (age and race), clinical keywords (UMLS concept unique identifiers from obstetric notes), clinical labs, and genetic risk (polygenic risk score for preterm birth). We compared three models for each feature set: 1) using only the feature set being evaluated (pink), 2) using only billing codes (‘Billing codes’, purple), and 3) using the feature set combined with billing codes (‘Both’, gray). For each feature set, we considered the subset of women who had at least one recorded value for the EHR feature and billing codes. All three models for a given EHR feature set considered the same pregnancies, but there are differences in the cohorts considered across features set due to differences in data availability; n_total_ is the number of women (training and held-out) for each feature set. **(B)** Each of the three models (x-axis) and their ROC-AUC and PR-AUC (y-axes) are shown. Each of the additional EHR features performed worse than the billing codes only model and did not substantially improve performance when combined with the billing codes. Dotted lines represent chance of 0.5 for ROC-AUC and the preterm birth prevalence for PR-AUC. Even when including EHR features before and after delivery in this framework revealed the same pattern with no substantial improvement in predictive performance compared to the billing codes only model (Fig. S6).

Models using only demographic factors, clinical keywords, and genetic risk had ROC-AUC and PR-AUC similar to chance (Fig. 3B). Clinical labs had moderate predictive power with ROC-AUC of 0.63 and PR-AUC of 0.24 (Fig. 3B). Compared to models using only billing codes, adding additional feature sets did not substantially improve performance (Fig. 3B). We note that some features sets, such as clinical labs and genetic risk, were evaluated on held-out sets with small numbers of deliveries (180 and 92, respectively). However, even after increasing the sample size by including women who may have features either before or after delivery, we did not observe a consistent gain in performance compared to models trained using only billing codes (Fig. S6).

### Models using billing codes outperforms prediction from risk factors

Although there are well known risk factors for preterm birth, few validated risk calculators exist and even fewer are routinely implemented in clinical practice[42]. We evaluated how a prediction model incorporating only common risk factors associated with moderate to high risk for preterm birth compared to a model using billing codes, which captured a broad range of comorbidities, at 28 weeks of gestation. We included maternal and fetal risk factors that occurred before and during the pregnancy and across many organ systems[3,13,23,43], race[20], age at delivery[44–46], pre-gestational and gestational diabetes[47], sickle cell disease[48], fetal abnormalities[13], pre-pregnancy hypertension, gestational hypertension (including pre-eclampsia or eclampsia)[1,49], and cervical abnormalities[50] (Methods).

The billing-code-based model significantly outperformed a model trained with clinical risk factors at predicting preterm birth at 28 weeks of gestation (PR-AUC=0.40 vs. 0.25, ROC-AUC=0.75 vs. 0.65; Fig. 4B, C). The stronger performance of the billing-code-based classifier was true for women across the spectrum of comorbidity burden; it had higher precision across individuals with different numbers of risk factors. Performance peaked for individuals with 0 (precision=0.39) and 4+ (precision=0.46) risk factors, but we did not observe a trend between model performance and increasing number of clinical risk factors (Fig. 4D). This suggests that machine learning approaches incorporating a comprehensive clinical phenome can add value to predicting preterm birth.

**Figure 4.**
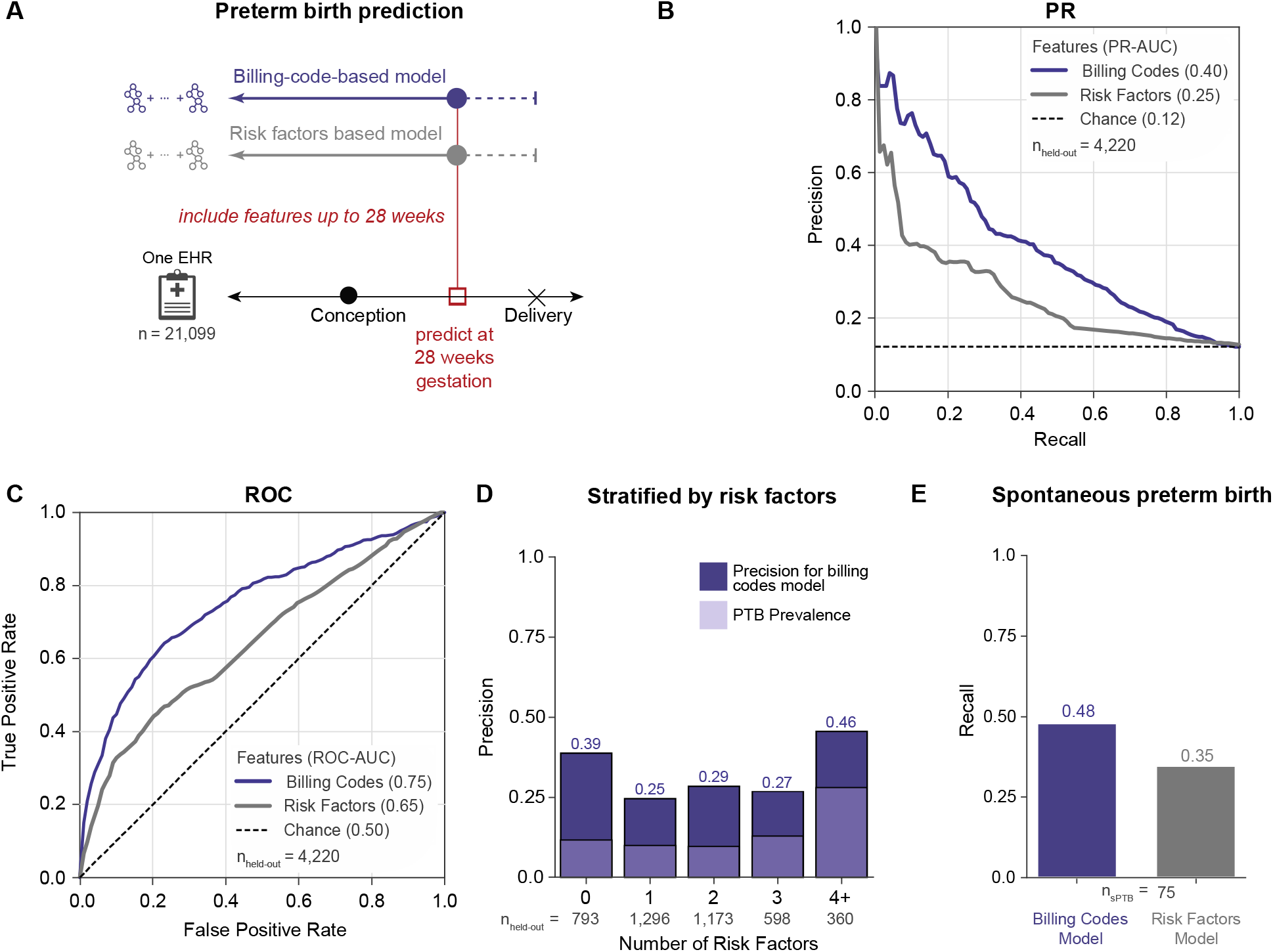
Billing-code-based model outperforms a model based on clinical risk factors. **(A)** We compared the performance of boosted decision trees trained using either billing codes (ICD-9 and CPT) present before 28 weeks of gestation (purple) or known clinical risk factors (gray) to predict preterm delivery. Clinical risk factors (Methods) included self- or third-party reported race (Black, Asian, or Hispanic), age at delivery (> 34 or <18 years old), non-gestational diabetes, gestational diabetes, sickle cell disease, presence of fetal abnormalities, pre-pregnancy BMI >35, pre-pregnancy hypertension (>120/80), gestational hypertension, preeclampsia, eclampsia, and cervical abnormalities. Both models were trained and evaluated on the same cohort of women (n = 21,099). **(B)** Precision-recall and **(C)** ROC curves for model using billing codes (purple line) or clinical risk factors (gray line). Preterm births are predicted more accurately by models using billing codes at 28 weeks of gestation (PR-AUC = 0.40, ROC-AUC = 0.75) than using clinical risk factors as features (PR-AUC = 0.25, ROC-AUC = 0.65). For the precision-recall curves chance performance is determined by the preterm birth prevalence (dashed black line). **(D)** Billing-code-based prediction model performance stratified by number of risk factors for an individual. The billing-code-based model detects more preterm cases and has higher precision (dark purple) across all numbers of risk factors compared to preterm (PTB) prevalence (light purple). **(E)** The model using billing codes also performs well at predicting the subset of spontaneous preterm births in the held-out set (recall = 0.48) compared to risk factors (recall = 0.35).

### Machine learning models can predict spontaneous preterm births

The multifactorial etiologies of preterm birth lead to clinical presentations with different comorbidities and trajectories. Medically-indicated and idiopathic spontaneous preterm births are distinct in etiologies and outcomes. Identifying pregnancies that ultimately result in spontaneous preterm deliveries is particularly valuable, and we anticipated that spontaneous preterm birth would be more challenging to predict than preterm birth overall. To test this, we identified spontaneous preterm births in the held-out set at 28 weeks of gestation by excluding women with medically induced labor, a cesarean section delivery, or PPROM (Methods). We intentionally used a conservative phenotyping strategy that aimed to minimize false positive spontaneous preterm births to evaluate the model’s ability to predict spontaneous preterm births. The prediction model trained using billing codes up to 28 weeks of gestation classified 48% (recall) of all spontaneous preterm births (n=75) as preterm; this is significantly higher than the risk factor only model (recall = 35%; Fig. 4E).

### Preterm birth prediction algorithm stratifies deliveries into clusters with different preterm birth risk and distinct comorbidity signatures

Understanding the statistical patterns identified by machine learning models is crucial for their adoption into clinical practice. Unlike deep learning approaches, decision tree-based models are easier to interpret. We calculated feature importance as measured by SHapley Additive exPlanation (SHAP) scores[51,52] for each delivery and feature pair in the held-out cohort for the model using billing codes at 28 weeks of gestation (‘Billing-code-based model’, Figure 4A). SHAP scores quantify the marginal additive contribution of each feature to the model predictions for each individual. Next, we performed a density-based clustering on the patient by feature importance matrix and visualized clusters using UMAP (Fig. 5A, Methods). This approach focuses the clustering on the features for each individual prioritized by the algorithm. We identified six clusters with 927 to 102 women. PTB prevalence was the high in the clusters one to four (blue, pink, green, orange, Fig. 5B) indicating differential risk for preterm birth. Performance varied across the clusters; the yellow cluster with low PTB prevalence had the highest PPV while clusters with higher PTB prevalence had higher recall (Fig. 5C, D).

**Figure 5.**
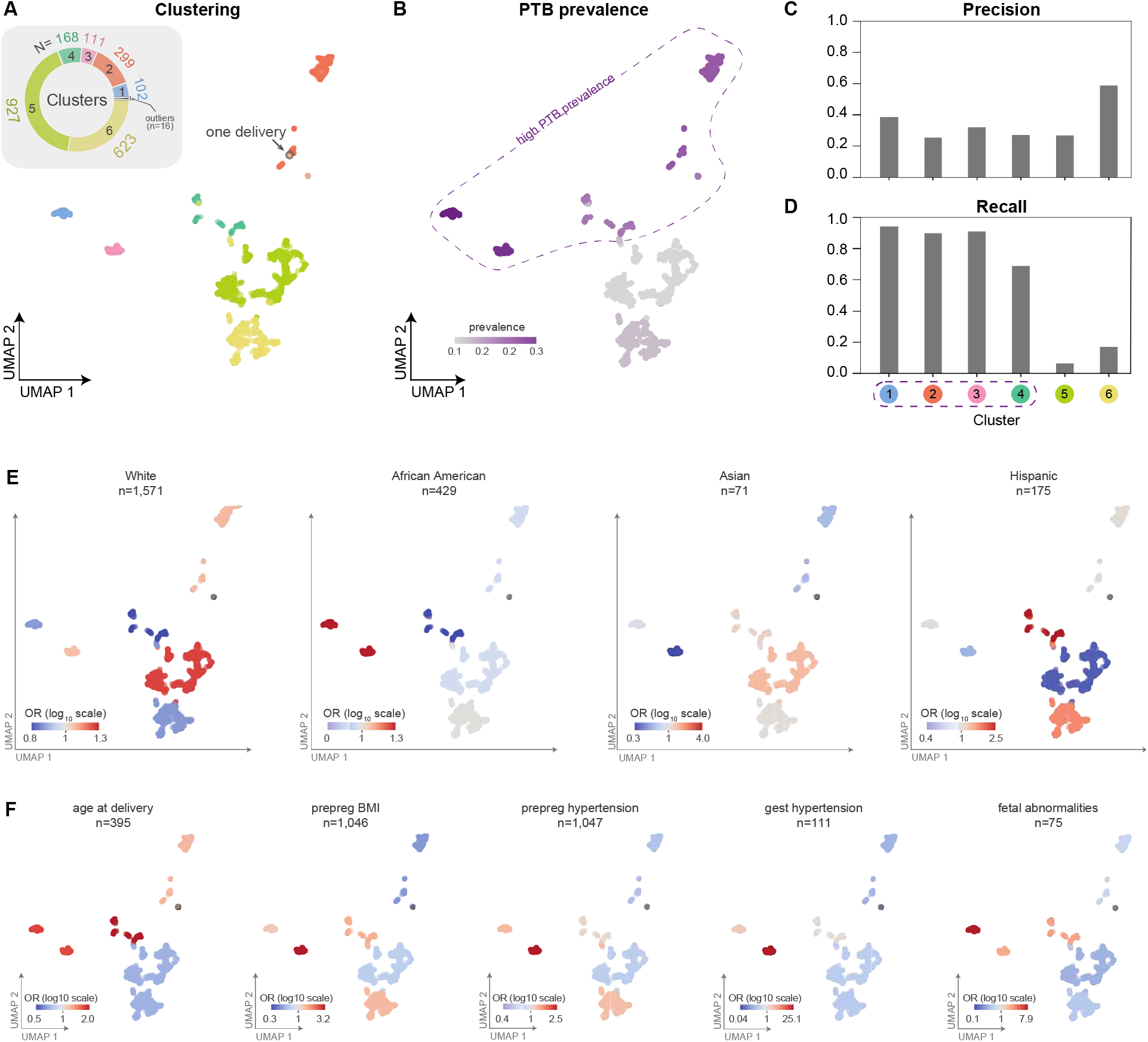
Machine-learning-based clustering of deliveries identifies sub-groups with distinct preterm birth prevalence, clinical features, and prediction accuracy. **(A)** For the model predicting preterm birth at 28 weeks of gestation using billing codes (ICD-9 and CPT, Figure 4A), we assigned deliveries from the held-out test set (n=2,246) to one of six clusters (colors) using density-based clustering (HDBSCAN) on the SHAP feature importance matrix. For visualization of the clusters, we used UMAP to embed the deliveries into a low dimensional space based on the matrix of feature importance values. Inset pie chart displays count of individuals in each cluster. **(B)** The preterm birth prevalence (colorbar) in each cluster. The algorithm discovered four clusters with high PTB prevalence (enclosed by dashed line). **(C)** Precision and **(D)** recall for preterm birth classification within each cluster. **(E)** The enrichment (odds ratios, colorbar in log10 scale) of race as derived from EHRs for each cluster (Table S1). **(F)** The enrichment (log_10_ odds ratio) of relevant clinical risk factors in each cluster (Table S2). Risk factors include: age at delivery (> 34 or <18 years old), pre-pregnancy BMI (prepreg BMI), pre-pregnancy hypertension (prepreg hypertension), gestational hypertension (gest hypertension), and fetal abnormalities. We report the total number of women in the delivery cohort at high risk for each clinical risk factor (n). Enrichments for additional risk factors are given in Fig. S7.

To evaluate whether clusters had distinct phenotype profiles, we calculated the enrichment of demographic and clinical risk factor traits within each cluster using Fisher’s exact test (Methods). These traits were extracted from structured fields in EHRs or ascertained using combinations of billing codes. Although these billing codes are used to train the model, the combination of codes used to ascertain risk factor traits are not encoded in the training data. White women are significantly enriched in the cluster 5 (odds ratio, OR = 1.2, p-value = 0.02, Fisher’s exact test, Figure 5E). Hispanic women also had significant positive enrichment in cluster four (OR = 2.5, p-value = 0.0002) and cluster 6 (OR = 1.6, p-value = 0.008) and were depleted (negative enrichment) in the cluster five (OR= 0.5, p-value = 4.42E-6, Figure 5D). African American and Asian women also exhibit modest enrichment in different clusters (Table S1).

We also tested for enrichment of clinical risk factors of preterm birth in the clusters. We observed distinct patterns of enrichment and depletion for each clinical risk factor (Fig. 5F, Fig. S7). Gestational hypertension had strong and enrichment in cluster three (OR = 26.4, p-value = 9.0E-39). Fetal abnormalities demonstrated a similar pattern with strong enrichment in cluster one (OR = 8.5, p-value = 2.07E-10). Extreme age at delivery (>34 or <18 years old) was enriched, though more weakly, (OR = 1.2 to 2.2) for the all clusters except five and six. Pre-pregnancy BMI, pre-pregnancy hypertension, and gestational hypertension had similar patterns with the strongest enrichment in cluster three. The remaining clinical risk factors show similar patterns and are provided in Fig. S7 and Table S2.

By analyzing the feature importance values through UMAP embeddings, we identify interpretable clusters of individuals discovered by the machine learning model that reflect the complex and multi-faceted paths to preterm birth. Overall, the learned rules highlight relationships between clinical factors and preterm birth prevalence. For example, some risk factors, such as age at delivery, are enriched in all clusters with high preterm birth prevalence. Other factors, such as pre-pregnancy BMI and hypertension, are strongly enriched only in specific clusters with high preterm birth prevalence. Thus, this approach enables us to interpret phenotypic patterns of risk and identify subgroups among cases learned from complex EHR features by the prediction model.

### Performance varies based on clinical context and delivery history

To further explore the sensitivity of the performance of our approach to clinical context and patient history, we evaluated how delivery type (vaginal vs. cesarean-section) and a previous preterm birth influence preterm birth prediction. We trained two classifiers using billing codes (ICD-9 and CPT) occurring before 28 weeks of gestation: one on a cohort of cesarean-section (n = 5,475) singleton deliveries and one on vaginal deliveries (n = 15,487). Preterm birth prediction accuracy was higher in the cesarean-section cohort (PR-AUC = 0.47, chance = 0.20) compared to the vaginal delivery cohort (PR-AUC = 0.23, chance = 0.10; Fig. 6A). Cesarean-sections also had higher ROC-AUC compared to vaginal deliveries (0.75 vs. 0.68, Fig. S8). As expected, the preterm birth prevalence was higher in the cesarean-section cohort.

**Figure 6.**
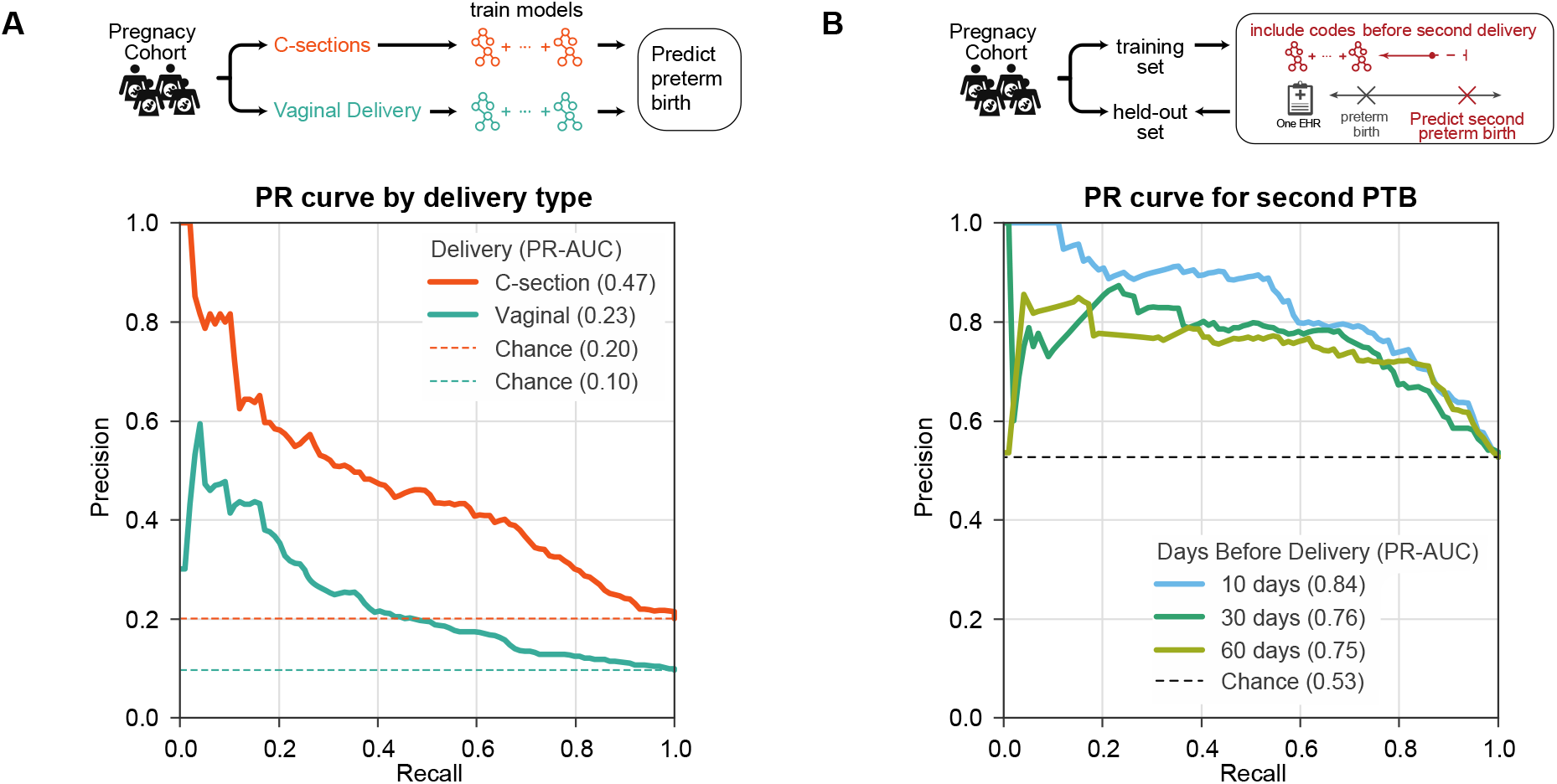
Preterm birth prediction accuracy is influenced by clinical context. **(A)** Preterm birth prediction models trained and evaluated only on cesarean section (C-section) deliveries perform better (PR-AUC=0.47) than those trained only on vaginal delivery (PR-AUC=0.23). ROC-AUC patterns were similar (Fig. S8). Billing codes (ICD-9 and CPT) present before 28 weeks of gestation were used to train a model to distinguish preterm from non-preterm birth for either C-sections (n=5,475) or vaginal deliveries (n=15,487). **(B)** Recurrent preterm birth can be accurately predicted from billing codes. We trained models to predict preterm birth for a second delivery in a cohort of 1,416 high-risk women with a prior preterm birth documented in their EHR. Three models were trained using data available at 10 days, 30 days, and 60 days before the date of second delivery. Models accurately predict the birth type in this cohort of women with a history of preterm birth (PR-AUC≥0.75). ROC-AUC varied from 0.82 at 10 days to 0.77 at 60 days before second delivery (Fig. S9). Expected performance by chance is the preterm birth prevalence in each cohort (dashed lines).

Women with a history of preterm birth are at significantly higher risk for a subsequent preterm birth than women without a previous history. Therefore, it is particularly important to understand the drivers of risk in this cohort. We tested if models trained on EHR data of women with a history of preterm birth could accurately predict the status of their next birth. We assembled 1,416 women with a preterm birth and a subsequent delivery in the cohort and split them into a training set (80%) and held-out test set (20%) to evaluate the model performance (Methods). For these women, 53% of the second deliveries were preterm. Due to limited availability of estimated gestational age data for the recurrent preterm births, which is necessary to approximate the date of conception, we trained models using billing codes (ICD-9 and CPT) present before each of the following timepoints: 10, 30, and 60 days before the delivery. These models were all able to discriminate term from preterm deliveries better than chance (Fig. 6B; PR-AUCs≥0.75). The model predicting a second preterm birth as early as 60 days before delivery achieved the high performance with PR-AUC=0.75 (Fig. 6B, chance=0.53) and ROC-AUC=0.77 (Fig. S9).

### Models trained at Vanderbilt accurately predict preterm birth in an independent cohort at UCSF

To evaluate whether preterm birth prediction models trained on the Vanderbilt cohort performed well on EHR data from other databases, we compared their performance on the held-out Vanderbilt cohort (n=4,215) and an independent cohort from UCSF (n=5,978). The UCSF cohort was ascertained using similar rules as the Vanderbilt cohort (Methods); age and distribution of race are provided in Table S3. However, we note that the UCSF cohort has a lower preterm birth prevalence (6%) compared to the Vanderbilt cohort (13%).

To facilitate the comparison, we trained models to predict preterm birth in the Vanderbilt cohort using only ICD-9 codes present before 28 weeks of gestation. We did not consider CPT codes in this analysis due to differences in the available billing code data between Vanderbilt and UCSF. As expected from the previous results, the model accurately predicted preterm birth in the held-out set from Vanderbilt (PR-AUC of 0.34, chance=0.12), but performance was slightly lower than using both ICD and CPT codes (Fig. 4B). The model trained at Vanderbilt also achieved strong performance in the UCSF cohort. The classifier had a higher ROC-AUC (0.80) in UCSF cohort compared to the Vanderbilt cohort (0.72; Fig. 7A) and PR-AUC of 0.31 vs 0.34 at Vanderbilt; Fig. 7B). The higher ROC is due to the lower prevalence of preterm birth in the UCSF cohort and the sensitivity of ROC-AUC to class imbalance[53]. Overall, these models show striking reproducibility across two independent cohorts.

**Figure 7.**
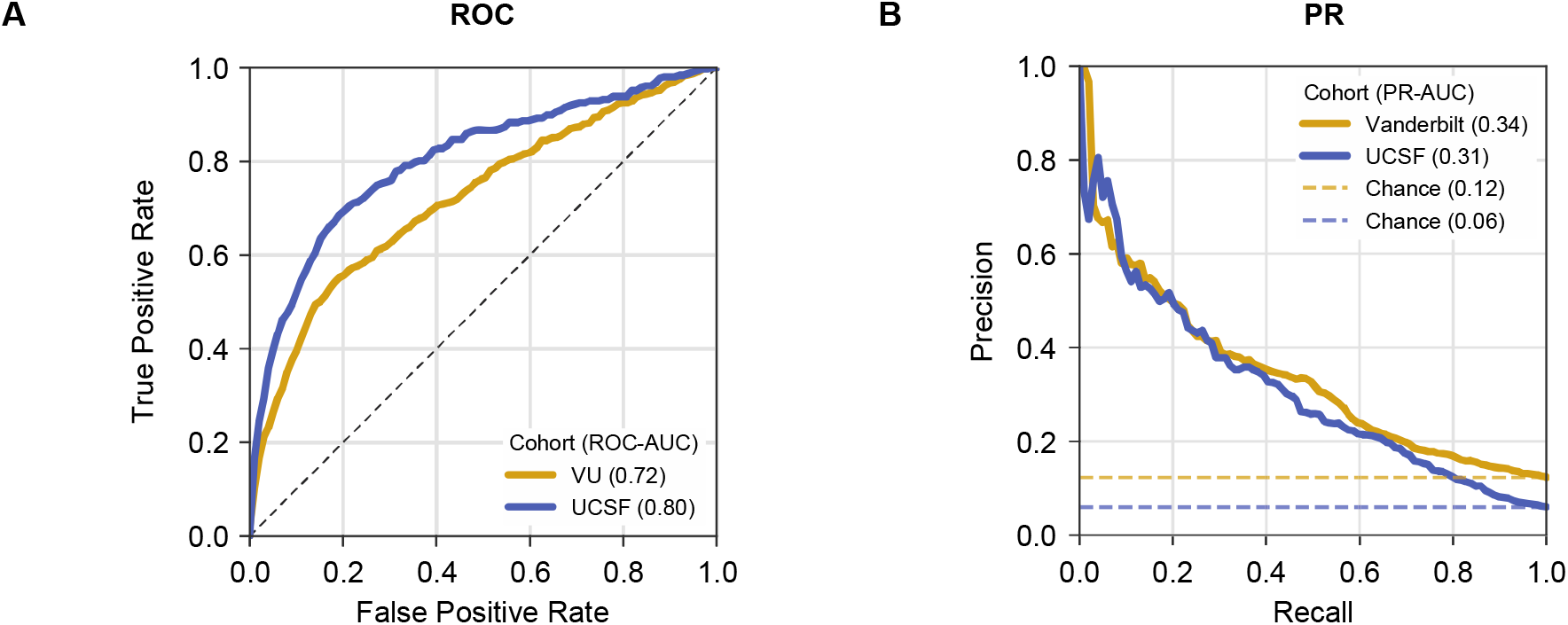
Preterm birth prediction models accurately generalize to an independent cohort. Performance of preterm birth prediction models trained at Vanderbilt applied to UCSF cohort. Models were trained using ICD-9 codes present before 28 weeks of gestation at Vanderbilt on 16,857 of women and evaluated on a held-out set at Vanderbilt (n=4,215, gold) and UCSF cohort (n=5,978, blue). (A) Models accurately predicted preterm birth at Vanderbilt (ROC-AUC=0.72) and at UCSF (ROC-AUC=0.80). The higher ROC-AUC at UCSF is driven by the lower prevalence of preterm birth in this cohort. (B) Models performed better than baseline prevalence (chance) based on the precision-recall curve at Vanderbilt (PR-AUC=0.34) and at UCSF (PR-AUC=0.31). Note that in contrast to models presented previously this one was trained only on ICD-9 codes, due to the lack of CPT codes in the UCSF cohort. Feature importance estimates were strongly correlated between the two cohorts (Fig. S10). Cohort demographics are given in Table S3.

### Similar features are predictive across the independent cohorts

The architecture of boosted decision trees enables straightforward identification of features (ICD-9 codes) with the largest influence on the model predictions. We used SHAP[54,55] scores to quantify the marginal additive contribution of each feature to the model predictions for each individual. For each feature in the ICD-9-based model, we calculated the mean absolute SHAP values across all women in the held-out set. The mean absolute SHAP value for each feature was highly correlated (spearman R=0.93, p-value < 2.2E-308) between the held-out Vanderbilt set and the UCSF cohort (Fig. S10A). The top 15 features ranked based on the mean absolute SHAP value captured known risk factors (fetal abnormalities, history of preterm birth, etc.), pregnancy screening and supervision of high-risk pregnancies (Fig. S10B). Ten of the top 15 features were shared across both cohorts. The full list of SHAP values across all features are provided in Table S4. This suggests that the model’s discovered combination of phenotypes, including expected risk factors, and the corresponding weights assigned by the machine learning model are generalizable across cohorts.

## Discussion

Preterm birth is a major health challenge affecting 5-20% of pregnancies[1,2,12] and lead to significant morbidity and mortality[56,57]. Predicting preterm birth risk could inform clinical management, but no accurate classification strategies are routinely implemented[24]. Here, we take a step toward addressing this need by demonstrating the potential for machine learning on dense phenotyping from EHRs to predict preterm birth in challenging clinical contexts (e.g., spontaneous and recurrent preterm births). However, we emphasize that more work is needed before these approaches are ready for the clinic. Compared to other data types in the EHRs, models using billing codes alone had the highest prediction accuracy and outperformed those using clinical risk factors. Demonstrating the potential broad applicability of our approach, the model accuracy was similar in an external independent cohort. Combinations of many known risk factors and patterns of care drove prediction; this suggests that the algorithm builds on existing knowledge. Thus, we conclude that machine learning based on EHR data has the potential to predict preterm birth accurately across multiple healthcare systems.

Decision tree based models are robust to correlated features, can identify complex non-linear combinations, and remain transparent for interpretation after training. In addition to these advantages, decision tree based models have demonstrated strong performance in various clinical prediction tasks[58–60]. Pregnancy is a clinical context with close monitoring and well defined end-points that may similarly benefit from machine learning approaches, yet few studies have applied decision tree based machine learning models to large pregnancy cohorts with rich clinical data[61].

Our approach has several distinct advantages compared to published preterm birth prediction models. First, our models have robust performance. Previous models using risk factors (diabetes, hypertension, sickle cell disease, history of preterm birth) to predict preterm birth, despite having cohorts up to two million women[23], have reported ROC-AUCs between 0.69 and 0.74[20–22]. Our models obtain a ROC-AUC of 0.75 and PR-AUC of 0.40 using data available at 28 weeks of gestation even after excluding multiple gestations. Furthermore, given the unbalanced classification problem (preterm births are less common than non-preterm), we report high PR-AUCs in addition to high ROC-AUCs. A recent deep learning model trained using word embeddings from EHRs achieved a high performance (ROC-AUC = 0.83[61]). This model was evaluated over a stratified high-risk cohort consisting of birth before 28 weeks of gestation. We did not stratify preterm births by severity since more than 85% of preterm births occur after 32 weeks of gestation[62], however, this is an important topic for future work. Our models achieve comparable performance with the benefit of easier interpretability, which is an advantage over deep learning approaches, and we discuss this further below.

Second, our models use readily available data throughout pregnancy that do not require invasive sampling. While some studies have also obtained high ROC-AUCs (e.g., 0.81-0.88), they used serum biomarkers across small cohorts[17] or acute obstetric changes within days of delivery[16]. The potential to enable cost-effective and broad application is illustrated by our evaluation of the classifiers on EHR data from UCSF; however, substantial further work is needed to move from this proof-of-concept analysis to clinic-ready models. Furthermore, the rich characterization of the phenome provided by EHRs leveraged by our approach could also complement more invasive biochemical assays.

Third, the gradient boosted decision trees we implement are easier to interpret than ‘black-box’ deep learning models that cannot easily identify features driving predictions. Transparency is an important, if not necessary, characteristic of machine/artificial learning models deployed in clinical practice[63,64], and it can facilitate discovery of insights and hypotheses to motivate future work. We reveal the patterns learned by our model by clustering deliveries using feature importance profiles. The enrichment for known risk factors in clusters with high preterm birth prevalence establishes confidence in our machine-learning based prediction models. In addition, we can quantify the strength of enrichment and combination of risk factors across clusters with distinct comorbid patterns. Since preterm birth is a heterogenous phenotype[6], and stratifying pregnancies based on clinical features may be critical to uncovering the biological basis of labor[3,65,66], the learned rules from our model offer a possible method for sub-phenotyping.

Finally, our approach generalizes across hospital systems. We demonstrate that billing-code-based models trained at Vanderbilt achieve similar accuracy in an independent cohort from UCSF. The generalizability of machine learning models can be constrained by the sampling of the training data. Thus, the accurate prediction in an independent dataset from an external institution points to several inherent strengths of the approach. First, successful replication indicates the models’ ability to learn predictive signals despite regional variation in assigning billing codes to an EHR. Second, the large cohorts used to train and evaluate models at Vanderbilt and USCF guard against potential weakness of EHRs, such as miscoding or omission of key data points. These errors are unavoidable in EHRs[67], but the large cohort used to train our models mitigates these errors and enables the high accuracy in the UCSF dataset, even with its different demographics. Additionally, idiosyncratic patterns of patient care at the institution used to develop the algorithm, which would be present in the Vanderbilt training and held-out sets, are unlikely to be present in the external UCSF cohort and inflate the out-of-sample accuracy. Third, the top features driving model performance are shared across institutions and reflect combinations of known risk factors and patterns of care. This aids interpretability of the underlying algorithm and likely reflects underlying pathophysiology that is innate to preterm birth.

We see several avenues for further improving our algorithm. First, some of the top features reflected routine obstetric care for high-risk pregnancies. Thus, the learning problem could be engineered to force the algorithm to discover new unappreciated risk factors. Second, we were surprised that the addition of features beyond billing codes, such as lab values, concepts extracted from clinical notes, and genetic information did not significantly improve performance. In some cases, any redundant information already captured by the billing codes would not improve the model’s accuracy; this is likely true for clinical notes. However, other sources, like currently available genetic data and polygenic risk scores, may not effectively capture underlying etiologies of preterm birth. Thus, these sources may not add more discriminatory power due to limitations in current data. Indeed, the largest published genome-wide study for preterm birth only explains a very small fraction of the heritability[30], and a polygenic risk score derived from it was not predictive in our cohort. Further sub-phenotyping of preterm birth will not only aid in prediction, but also understanding its multifactorial etiology and developing personalized treatment strategies. More explicit modeling of the temporal dependence between EHR features may further increase performance. Finally, while we evaluated the ability of our classifiers to discriminate preterm births, further studies evaluating the calibration of these models are necessary to better risk stratify of pregnancies.

The strong predictive performance of our models suggests that they have the potential to be clinically useful. Compared to a machine learning model trained using only known risk factors, the billing-code-based classifier incorporated a broad set of clinical features and predicted preterm birth with higher accuracy. Furthermore, the superior performance was not driven by the number of risk factors or the total burden of billing codes. These results indicate the algorithm is not simply identifying less healthy individuals or those with greater healthcare usage. The models also accurately predicted many preterm births in challenging and important clinical contexts such as spontaneous and recurrent preterm birth. Spontaneous preterm births are common[1,12,68], and unlike iatrogenic deliveries, they are more difficult to predict because they are driven by unknown multifactorial etiologies[12,24]. Similarly, since a prior history of preterm birth is one of the strongest risk factors[69], distinguishing pregnancies most at risk for recurrent preterm birth has potential to provide clinical value.

However, we emphasize that additional work is needed before this approach is ready for clinical application. Though it has strong performance, a more comprehensive evaluation of the algorithm against current clinical practice is needed to determine how early and how much improvement in standard of care this approach could provide[70]. Furthermore, while our cohorts include diverse individuals and the algorithm generalizes well, the approach must be evaluated to ensure that it does not introduce of amplify biases against specific groups or types of preterm birth[71]. In addition, we anticipate further gains in the clinical value of this approach as more modalities of data becomes incorporated in the EHR[72] and diverse populations become available. Addressing these questions and taking other necessary steps toward clinical utility will require the close collaboration of diverse experts from basic, clinical, social, and implementation sciences.

Our results provide a proof-of-concept that machine learning algorithms can use the dense phenotype information collected during pregnancy in EHRs to predict preterm birth. The prediction accuracy across clinical contexts and compared to existing risk factors suggests such modeling strategies can be clinically useful. We are optimistic that with the increasingly widespread adpotion of, improvement in tools for extracting meaningful data from them, and integration of complementary molecular data, machine learning approaches can improve the clinical management of preterm birth.

## Materials and Methods

### Ascertaining delivery type and date for Vanderbilt cohort

We identified women with at least one delivery (n=35,282, ‘delivery-cohort’) at Vanderbilt Hospital based on the presence of delivery-specific billing codes (ICD-9/10 and CPT) or estimated gestational age (EGA) documented in the EHR. Combining delivery specific ICD-9/10 (‘delivery-ICDs’), CPT (‘delivery-CPTs’), and EGA data, we developed an algorithm to label each delivery as preterm or not preterm. Women with multiple gestations (e.g. twins, triplets) were identified using ICD and CPT codes and exclude for singleton-based analyses. See Supplementary Materials and Methods for exact codes.

We demarcate multiple deliveries by grouping delivery-ICDs in intervals of 37 weeks starting with the most recent delivery-ICD. This step is repeated until all delivery-ICDs in a patient’s EHR are assigned to a pregnancy. We chose 37-week intervals to maximally discriminate between pregnancies. For each delivery, we assign a list of labels (preterm, term, or postterm) ascertained using the delivery-ICDs. EGA values, extracted from structured fields across clinical notes, were mapped to multiple pregnancies using the same procedure. For women with multiple EGA recorded in their EHR, the most recent EGA value determined the time interval to group preceding EGA values. Based on the most recent EGA value for each pregnancy, we assigned labels to each delivery (EGA <37 weeks: preterm; ≥37 and <42 weeks: term, ≥42 weeks: postterm). After pooling delivery labels based on delivery-ICDs and EGA, we assigned a consensus delivery label by selecting the oldest gestational age based classification (i.e. postterm > term > preterm). By incorporating both billing code and EGA based delivery label and selecting the oldest gestational classification, we expect this to increase the accuracy of this algorithm, which we evaluate by chart-review (described in detail below).

Since CPT codes do not encode delivery type, we combined the delivery-CPTs with timestamps of delivery-ICDs and EGAs to approximate the date of delivery. Delivery-CPTs were grouped into multiple pregnancies as described above. The most recent timestamp from delivery-CPTs, delivery-ICDs, and EGA values was used as the approximate delivery date for a given pregnancy.

### Validating delivery type based on chart review

To validate the delivery type ascertained from billing codes and EGA, we used chart-reviewed labels as the gold standard. For 104 randomly selected EHRs from the delivery cohort, we extracted the date and gestational age at delivery from clinical notes. For earliest delivery recorded in the EHR, we assigned a chart-review based label according to the gestational age at delivery (<37 weeks: preterm; 37 and 42 weeks: term, ≥42 weeks: postterm). The precision/positive predictive value for the ascertained delivery type as a binary variable (‘preterm’ or ‘not-preterm’) was calculated using the chart reviewed label as the gold standard. To compare the ascertainment strategy to a simpler phenotyping algorithm, we compared the concordance of the label derived from delivery-ICDs to one based on the gestational age within three days of delivery. This simpler phenotyping approach resulted in a lower PPV (85%) and recall (93%; Fig. S1B) compared to the billing-code-based ascertainment strategy.

### Training and evaluating gradient boosted decision trees to predict preterm birth

All models for predicting preterm birth used boosted decision trees as implemented in XGBoost v0.82[38]. Unless stated otherwise, we trained models to predict the earliest delivery in a woman’s EHR as preterm or not-preterm. The delivery cohort was randomly split into training (80%) and held-out (20%) sets with equal proportion of preterm cases. For prediction tasks, we used only ICD-9 and excluded ICD-10 codes to avoid potential confounding effects. The total count of billing codes within a specified time frame was used as features to train our models; if a woman never had a billing code in her EHR, we encoded these as ‘0’. For all models we excluded ICD-9, CPT codes, and EGA used to ascertain delivery type and date. On the training set, we use tree of Parzen estimators as implemented in hyperopt v0.1.1[73] to optimize hyperparameters by maximizing the mean average precision. The best set of hyperparameters was selected after 1,000 trials using 3-fold cross-validation over the training set (80:20 split with equal proportion of preterm cases). We evaluated the performance of all models on the held-out set using Scikit-learn v0.20.2[74]. All performance metrics reported are on the held-out set. For precision-recall curves, we define baseline chance for each model as the prevalence of preterm cases. To ensure no data leaks were present in our training protocol, we trained and evaluated a model using a randomly generated dataset (n=1,000 samples) with a 22% preterm prevalence. As expected, this model did not do better than chance (AUC=0.50, PR-AUC=0.22, data not show). All trained models with their optimized hyperparameters are provided at https://github.com/abraham-abin13/ptb_predict_ml.

### Predicting preterm birth at different weeks of gestation

As a first step, we evaluated whether billing codes could discriminate between delivery types. Models were trained to predict preterm birth using the total counts of each ICD-9, CPT, or ICD-9 and CPT code across a woman’s EHR. We excluded any codes used to ascertain delivery type or date. All three models were trained and evaluated on the same cohort of women who had at least one ICD-9 and CPT code (Fig S2).

Next, we evaluated machine learning models at 0, 13, 28, and 35 weeks of gestation by training using only features present before each timepoint. For the subset of women in our delivery cohort with EGA, we calculated the date of conception by subtracting EGA (recorded within three days of delivery) from the date of delivery. Next, we trained models using ICD-9 and CPT codes timestamped before different gestational timepoints with only singleton (Fig. 2B) or including multiple gestations (Fig. S3). The same cohort of women was used to train and evaluate across models. The sample size varied slightly (n = 11,843 to 10,799) since women who already delivered were excluded at each timepoint.

In addition to evaluating models based on the date of conception, we trained models at different timepoints before the date of delivery (Fig. S4) using the same cohort of women by requiring every individual in this cohort had to have at least one ICD-9 or CPT code before each timepoint. Evaluating models before the date of delivery increased the sample size (n=15,481) compared to a prospective conception-based design (n=12,410) and yielded similar results.

### Evaluating predictive potential of demographic, clinical, and genetic features from EHRs

In addition to billing codes, we extracted structured and unstructured features from the EHRs (Fig. 3A). We evaluated models using features present before 28 weeks of gestations (Fig 3) and features present before or after delivery (Fig S6). Structured data included self or third-party reported race (Fig. 1E), age at delivery, past medical and family history (92 features, see Supplementary Materials and Methods), and clinical labs. For training models, we only included clinical labs obtained during the first pregnancy and excluded values greater than four standard deviations from the mean. To capture the trajectory of each clinical lab’s values across pregnancy (307 clinical labs, see Supplementary Materials and Methods), we trained models using the mean, median, minimum, and maximum lab measurement. For unstructured clinical text in obstetric and nursing clinical notes, we applied CLAMP[75] to extract UMLS (Unified Medical Language System) concepts unique identifiers (CUIs and included those with positive assertions with > 0.5% frequency across all EHRs). When training preterm birth prediction models, we one-hot encoded categorical features. No transformations were applied to the continuous features.

A subset of women (n=905) was genotyped on the Illumina MEGA^EX^ platform. We applied standard GWAS quality control steps[76] using PLINK v1.90b4s[77]. We calculated a polygenic risk score for each white woman with genotype data based on the largest available preterm birth GWAS [30] using PRSice-2[78,79]. We assumed an additive model and summed the number of risk alleles at single nucleotide polymorphisms (SNPs) weighted by their strength of association with preterm birth (effect size). PRSice determined the optimum number of SNPs by testing the polygenic risk score for association with preterm birth in our delivery-cohort at different GWAS p-value thresholds. We included date of birth and five genetic principal components to control for ancestry. Our final polygenic risk score used 356 preterm birth associated SNPs (GWAS p-value < 0.00025).

Using the structured and unstructured data derived from the EHR, we evaluated whether adding EHR features to billing codes could improve preterm birth prediction. Since the number of women varied across EHR feature, we created subsets of the delivery cohort for each EHR feature. Each subset included women with at least one recorded value for the EHR feature and billing codes. Then we trained three models as described above for each subset: 1) using only the EHR feature being evaluated, 2) using ICD-9 & CPT codes, and 3) using the EHR feature with ICD-9 & CPT codes. Thus, all three models for a given EHR feature were trained and evaluated on the same cohort of deliveries (Fig. 3A).

### Predicting preterm birth using billing codes and clinical risk factors at 28 weeks of gestation

We compared the performance of a model trained using billing codes (ICD-9 and CPT) present before 28 weeks of gestation with a model trained using clinical risk factors to predict preterm delivery (Fig 4). Both models were trained and evaluated on the same cohort of women (n = 21,099). We selected well-established obstetric risk factors that included maternal and fetal factors across organ systems, occurred before and during pregnancy, and had moderate to high risk for preterm birth [3,13,23,43]. For each individual, risk factors were encoded as high-risk or low-risk binary values. Risk factors such as non-gestational diabetes status[47], gestational diabetes[47], gestational hypertension, pre-eclampsia or eclampsia[1,49], fetal abnormalities[13], cervical abnormalities[50], and sickle cell disease[48] status was defined based on at least one corresponding ICD-9 code occurring before the date of delivery (Supplementary Materials and Methods). The remaining factors, such as race (Black, Asian, or Hispanic was encoded as higher risk)[20], age at delivery (> 34 or <18 years old)[44–46], pre-pregnancy BMI ≥ 35, and pre-pregnancy hypertension (>120/80)[1,49], were extracted from structured fields in EHR. Pre-pregnancy value was defined as the most recent measurement occurring before nine months of the delivery date.

### Density based clustering on feature importance values

To better understand the decision making process of our machine learning models, we calculated feature importance value for the model predicting preterm birth at 28 weeks of gestation. We used SHapley Additive exPlanation values (SHAP)[51,52,55] to determine the marginal additive contribution of each feature for each individual. First, we calculated a matrix of SHAP values of features by individuals from the held-out cohort. Since the shape of this matrix was too large to perform the density based clustering, we created an embedding using 30 UMAP components with default parameters as implemented in UMAPv0.3.8[80]. Next, we performed a density based hierarchical clustering using HDBSCANv0.8.26 [81]. We used default parameters (metric=Euclidean) and tried a range of values for two hyperparameters: minimum number of individuals in each cluster (‘min_clust_size) and threshold for determining outlier individuals who do not belong to a cluster (‘min_samples’). After tuning these two hyperparameters, we selected the clustering model with the highest density based cluster validity score [81], which measures the within and between cluster density connectedness. We find a min_clust_size = 110 and min_samples = 10 had the highest density based cluster validity (DBCV) score with 6 distinct clusters with one cluster for outliers (Fig. S11). A minority of women (n=16) were not assigned to a cluster (‘outliers’). To visualize the cluster assignments, we performed UMAP on the feature importance matrix with default settings and two UMAP components and colored each individual by their cluster membership. Finally, we calculated the preterm birth prevalence and accuracy within each cluster.

### Comorbidity enrichment within clusters

We tested for enrichment of clinical risk factors within each cluster by using a Fisher Exact test as implemented in Scipy[82]. For each risk factor, we constructed a contingency table based on a given cluster membership and being high risk for the risk factor. We report enrichment as the odds ratio with colorbar in log10 scale of the odds ratio. For sickle cell disease, one cluster did not have any cases of sickle cell disease.

### Evaluating model performance on spontaneous preterm births, by delivery type, and recurrent preterm birth

We compared how models trained used billing codes (ICD-9 & CPT) performed in different clinical contexts. First, we evaluated the accuracy of predicting spontaneous preterm birth using models trained to predict all types of preterm births. From all preterm cases in the held-out set, we excluded women who met any of the following criteria to create a cohort of spontaneous preterm births: medically induced labor, delivery by cesarean section, or preterm premature rupture of membranes. The ICD-9 and CPT codes used to identify exclusion criteria are provided in Supplementary Materials and Methods. We calculated recall/sensitivity as the number of predicted spontaneous preterm births out of all spontaneous preterm births in the held-out set. We used the same approach to quantify performance of models trained using clinical risk factors (Fig. 4E).

We trained models to predict preterm birth among cesarean sections and vaginal deliveries separately using billing codes (ICD-9 & CPT) as features. Deliveries were labeled as cesarean sections or vaginal deliveries if they had at least one relevant billing code (ICD-9 or CPT) occurring within ten days of the date of first delivery in EHR. Billing codes used to determine delivery type are provided in Supplementary Materials and Methods. Deliveries with billing codes for both cesarean and vaginal deliveries were excluded. We trained separate models to predict cesarean and vaginal deliveries (Fig. 6A and Fig. S8).

We evaluated how well models using billing codes could predict recurrent preterm birth. From our delivery cohort, we retained women whose first delivery in the EHR was preterm and a second delivery for which we ascertained the type (preterm vs. not-preterm) as described above for the first delivery. We trained models using billing codes (ICD-9 & CPT) at timepoints before the date of delivery because the majority of this cohort did not have reliable EGA at the second delivery. As described earlier, separate models were trained using billing codes timestamped before timepoint being evaluated (Fig. 6B, Fig. S9).

### Preterm birth prediction in independent UCSF cohort

We evaluated how well models trained at Vanderbilt using billing codes would replicate in an external cohort assembled at UCSF. Only the first delivery in the EHR was used for prediction. Women with twins or multiple gestations, identified using billing codes (Supplementary Materials and Methods), were excluded. Delivery type (preterm vs. not preterm) was assigned based on the presence of ICD-10 codes. Term (or not-preterm) deliveries were determined by the presence of an ICD-10 code beginning with the characters “O80”, specifying an encounter for full-term delivery. Preterm deliveries were determined by both the absence of ICD-10 codes beginning with “O80” and the presence of codes beginning with “O60.1”, the family of codes for preterm labor with preterm delivery. We trained models using ICD-9 codes present before 28 weeks of gestation on the Vanderbilt cohort to predict preterm birth. CPT codes were not used since they were not available from the UCSF EHR system. The 28-week model was evaluated on the Vanderbilt held-out set and the independent UCSF cohort.

### Feature interpretation from boosted decision tree models

To determine feature importance, we used SHapley Additive exPlanation values (SHAP)[52,54,55] to determine the marginal additive contribution of each feature. For the held-out Vanderbilt cohort and the UCSF cohort, a SHAP value was calculated for each feature per individual. Feature importance was summarized by taking the mean of the absolute value of SHAP scores across individuals. The top fifteen features based on the mean absolute SHAP value in either the Vanderbilt or UCSF cohorts values are reported. To compare how feature importance varies at Vanderbilt and UCSF, we computed the Pearson correlation of the mean absolute SHAP values.

## Data Availability

All code and models in this study are available at https://github.com/abraham-abin13/ptb_predict_ml.

## Ethics

This study exclusively utilized information extracted from medical records in the Vanderbilt University Medical Center (VUMC) “Synthetic Derivative” database (SD). The SD is a de-identified copy of the main hospital medical record databases created for research purposes. The de-identification of SD records was achieved primarily through the application of a commercial electronic program, which was applied and assessed for acceptable effectiveness in scrubbing identifiers. For instance, if the name “John Smith” appeared in the original medical record, its corresponding record in the SD does not contain “John Smith”. Instead, it is permanently replaced with a tag [NAMEAAA, BBB] to maintain the semantic integrity of the text. Similarly, dates, such as “January 1, 2004” have been replaced with a randomly generated date, such as “February 3, 2003.”

The SD database (which contains over 3 million electronic records, with no defined exclusions) was accessed through database queries. Searches are logged and audited annually. As no HIPAA identifiers are available in the SD database, and this work does not plan to re-identify these records using the identified VUMC database, this study meets criteria for non-human subjects research. Nonetheless, to ensure confidentiality and appropriate use of the SD, all relevant key personnel for this study entered a data use agreement, which prohibits any use of the data not described in this application, including the re-identification of the SD records.

## Acknowledgments

We thank the members of the Capra lab and the March of Dimes Prematurity Research Center Ohio Collaborative for thoughtful discussion on this project.

## Funding

AA was supported by the American Heart Association fellowship 20PRE35080073, National Institutes of Health (NIH, T32GM007347), the March of Dimes, and the Burroughs Wellcome Fund. MS, IK, and BL were supported by the March of Dimes and NIH (NLM K01LM012381). JAC was supported by the NIH (R35GM127087), NIH (1R01HD101669), March of Dimes, and the Burroughs Wellcome Fund. This work was conducted in part using the resources of the Advanced Computing Center for Research and Education at Vanderbilt University. The dataset(s) used for the analyses described were obtained from Vanderbilt University Medical Center’s BioVU which is supported by numerous sources: institutional funding, private agencies, and federal grants. These include the NIH funded Shared Instrumentation Grant S10RR025141; and CTSA grants UL1TR002243, UL1TR000445, and UL1RR024975. Genomic data are also supported by investigator-led projects that include U01HG004798, R01NS032830, RC2GM092618, P50GM115305, U01HG006378, U19HL065962, R01HD074711; and additional funding sources listed at https://victr.vumc.org/biovu-funding/. The content is solely the responsibility of the authors and does not necessarily represent the official views of the National Institutes of Health, the March of Dimes, or the Burroughs Wellcome Fund.

## Author contributions

*Conceptualization, Methodology:* A.A. and J.A.C. conceived and designed the study. J.M.N provided clincial interpretation and aided in feature selection. *Data curation:* A.A. and C.A.B. extracted billing codes, clinical notes, and performed concept extraction on the Vanderbilt cohort. A.A., P.S. and L.K.D. extracted, cleaned, and provided clinical laboratory data during pregnancy on the Vanderbilt cohort. *Resources:* D.R.V. provided obstetric and nursing notes on the Vanderbilt cohort. B.L., I.K., M.S. extracted the delivery cohort from UCSF. *Formal Analysis, Investigation:* A.A. performed all analyses on the Vanderbilt cohort under supervision from J.A.C. B.L. and I.K. evaluated models on UCSF cohorts under supervision from M.S. *Funding acquisition:* J.A.C. *Writing:* A.A. wrote the manuscript with guidance from J.A.C., J.M.N., M.S., L.M. and A.R.

## Competing interests

LJM is a consultant for Mirvie, Inc.

## Data and materials availability

The dataset(s) and code supporting the conclusions of this article is(are) available in the https://github.com/abraham-abin13/ptb_predict_ml repository.

## Notes

### Funding Statement

For funding, please see pdf of manuscript.

### Author Declarations

A designee of the Vanderbilt Institutional Review Board reviewed the research study identified above. The designee determined the study does not qualify as "human subject" research per Section 46.102(f)(2).

