## Supplement for "Dense phenotyping from electronic health records enables machine-learning-based prediction of preterm birth"

Supplemental Figures

Supplemental Tables

Supplemental Methods

**Supplemental Figures**

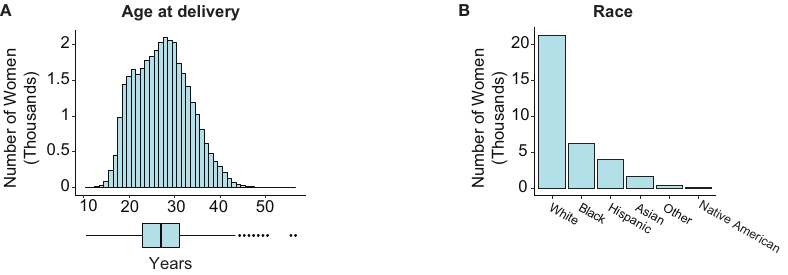

**Fig. S1.** **Distribution of maternal age at delivery and self- or third-party reported race. (A)** The distribution of age at first delivery in EHR (mean 27.3 years; 23.0–31.0 years, 25^th^ and 75^th^ percentiles). **(B)** Counts of women by self- or third-party reported race (White: 21,343; Black: 6,178; Hispanic: 3,979; Asian: 1,617; Other: 409; Native American: 84).

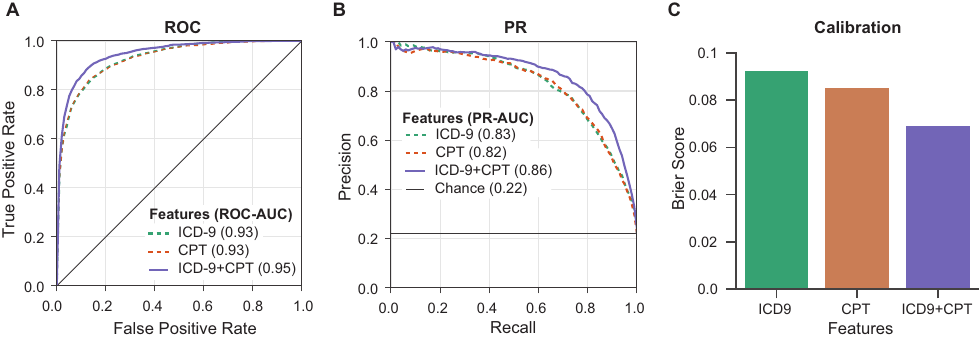

**Fig. S2: Boosted decision trees trained on EHR billing codes accurately identify preterm births.** We trained and validated boosted decision trees on 80% of labeled pregnancies (preterm vs. non-preterm) from the EHR cohort (n=35,282, Fig. 1). We included both singletons and multiple gestations. We evaluated model performance on the held-out set using area under the ROC and precision-recall curves (ROC-AUC, PR-AUC) and Brier scores. EHR features used to ascertain delivery labels are excluded in training and evaluation of the models. **(A,B)** The boosted decision trees accurately classified deliveries by preterm birth status using only ICD-9 (green dashed line), only CPT (orange dashed line), and combined ICD-9 and CPT (solid purple) features present in a women’s EHR, (ROC-AUC≥0.93, PR-AUC≥0.86). Combining ICD-9 and CPT codes achieved the best performance. **(C)** The low Brier scores (≤0.092) indicate that the models are well calibrated.

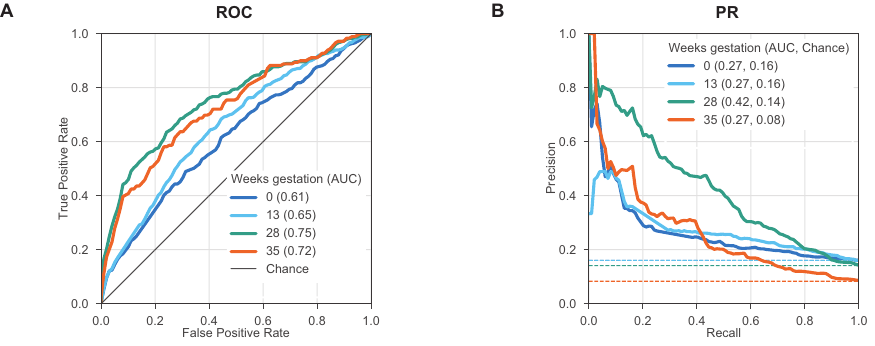
**Fig. S3: Machine learning can accurately identify preterm birth including singletons and multiple gestations.** We trained models (boosted decision trees) on 80% of the corresponding cohort to predict the earliest delivery as preterm or not-preterm (Methods). In contrast to the models presented in the main text (Fig. 2), these included singleton and multiple gestations. Billing codes (ICD-9 and CPT) present before pregnancy (0, 13, 28, and 35 week of gestation) were used to train models. The same cohort of women (training + held-out) was used to train and evaluate across models but the sample size varied slightly (n = 11,843 to 10,799) since women who already delivered were excluded at each timepoint. **(A)** The ROC-AUC increased from conception at 0 weeks (0.61, dark blue line) to 35 weeks of gestation (0.72, orange line) compared to a chance (black line). **(B)** The model at 28 weeks of gestation achieved the highest PR-AUC (0.42). Chance (dashed lines) represents the preterm birth prevalence in each cohort.

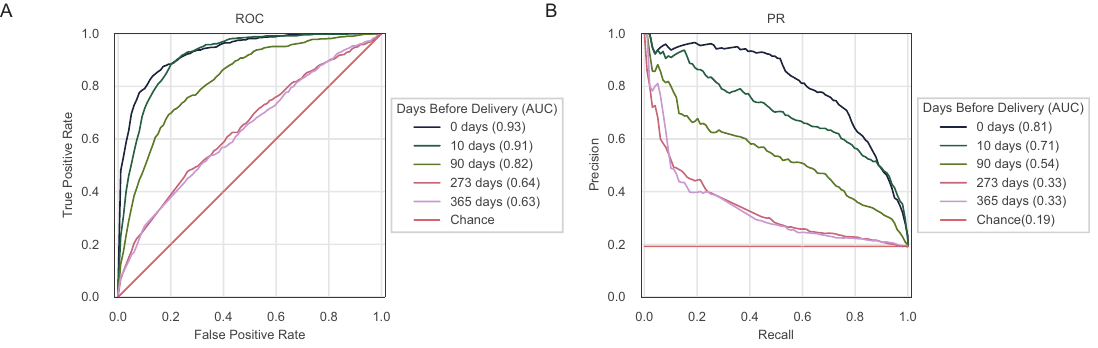

**Fig. S4:** **Preterm birth prediction increases at timepoints closer to the date of delivery at timepoints based on days before delivery.** **(A)** ROC and **(B)** PR curves for preterm birth prediction using billing codes (ICD-9 & CPT) at different timepoints defined from the date of delivery in the Vanderbilt cohort. Both singletons and multiple gestations are included. Chance for PR-AUC represent random prediction equal to the population prevalence of preterm birth. Model performance improves as the prediction is made closer to delivery. All models are trained and evaluated on the same cohort of women (n=15,481) and the performance reported is on the held-out set (20% of cohort).

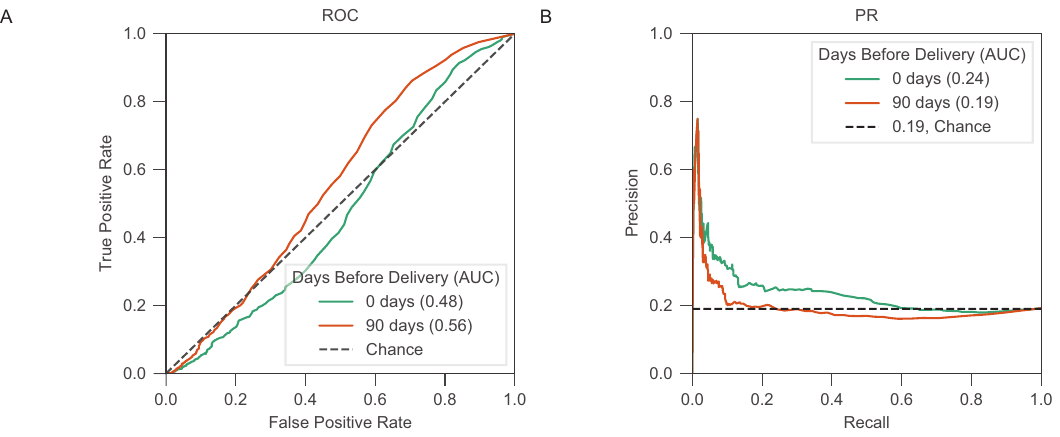
**Fig. S5: Preterm birth prediction is not driven by total number of billing codes.**

To evaluate whether the amount of contact with the healthcare system was driving the performance of our machine learning classifiers, we assessed the discriminatory ability of the total number of billing codes (ICD-9 or CPT) in a woman’s EHR to predict preterm birth. We include both singletons and multiple gestations. A simple classifier that used only the number of total billing codes preset at 0 days (green) and 90 days (orange) before the first delivery in her EHR, did not predict preterm birth well: **(A)** ROC-AUC = 0.56 and **(B)** PR-AUC = 0.19. The cohort consisted of the held-out set at the specified timepoints with 3,096 women.

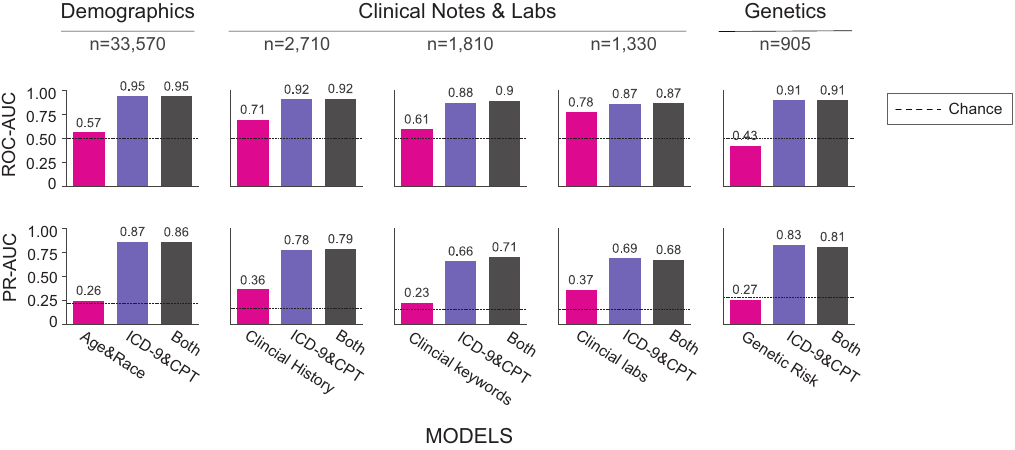

**Fig. S6: Combining EHR features with billing codes does not improve model performance.** We evaluated how combining EHR features with billing codes could improve model performance. We used a similar framework as stated in Figure 3 but included billing codes and features before and after delivery instead of before 28 weeks of gestation. We also included multiple gestations instead of only including singleton preganncies. EHR features are grouped in to sets of: demographic factors (age and race), clinical history (patient and familial comorbidities), clinical keywords (UMLS concept unique identifiers from obstetric notes), clinical labs, and genetic risk (polygenic risk score for preterm birth). We compared three models for each feature set: 1) using only the feature set being evaluated (pink), 2) using ICD-9 & CPT codes (purple), and 3) using the feature set combined with ICD-9&CPT codes (gray). For each feature set, we considered the subset of women who had at least one recorded value for the EHR feature and ICD&CPT codes. All three models for a given EHR feature set considered the same pregnancies, but there are differences in the cohorts considered across features set due to differences in data availability. Each of the three models (x-axis) and their ROC-AUC and PR-AUC (y-axes) are shown. Each of the additional EHR features performed worse than the billing code only model and did not substantially improve performance when combined with the billing codes. Of the other EHR features tested, clinical labs had the best predictive performance with PR-AUC of 0.37 and ROC-AUC of 0.78. Dotted lines represent chance of 0.5 for ROC-AUC and the preterm birth prevalence for PR-AUC. The total number of women (n) in each subset including the training and held-out set is given.

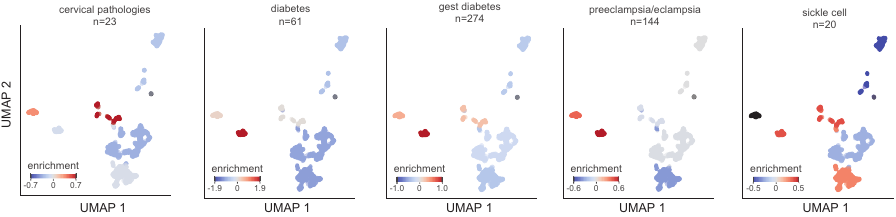

**Fig. S7: Enrichment of additional clinical risk factors in pregnancy cohort clusters.** We calculated enrichment **(**log_10_ odds ratio) of several additional clinical risk factors (each panel) for each cluster derived from the feature importance matrix for the model predicting preterm birth at 28 weeks of gestation (Figure 5, Methods). These risk factors are enriched in different clusters. We report the total number of women in the delivery cohort at high risk for each clinical risk factor (n).

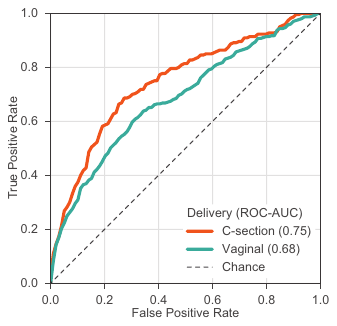

**Fig. S8: Preterm birth prediction accuracy is higher for cesarean-sections compared to vaginal deliveries.** After stratifying the delivery cohort into cesarean-sections (n=5,475) and vaginal (n=15,487) deliveries, we trained a model on each delivery type to predict preterm or not-preterm births. Multiple gestations were excluded. We trained models using billing codes (ICD-9 and CPT) present before 28 weeks of gestation. ROC-AUC was higher for cesarean-sections (0.75) compared to vaginal deliveries (0.68). This corresponds to the PR curves presented in Fig. 6A.

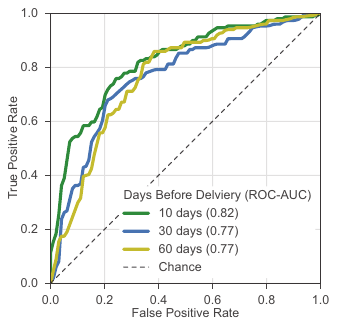

**Fig. S9: Models trained using billing codes can accurately predict risk of a second preterm birth.** For women with a history of preterm birth (n=1,416, Methods), we trained models using billing codes (ICD-9 and CPT) to predict a second preterm birth. Multiple gestations were excluded. For each model, only billing codes timestamped before the specified number of days before delivery are included. Models predicted a second preterm birth accurately with the highest and lowest ROC-AUC of 0.82 at 10 days and 0.77 at 60 days before delivery respectively. This corresponds to the PR curves presented in Fig. 6B.

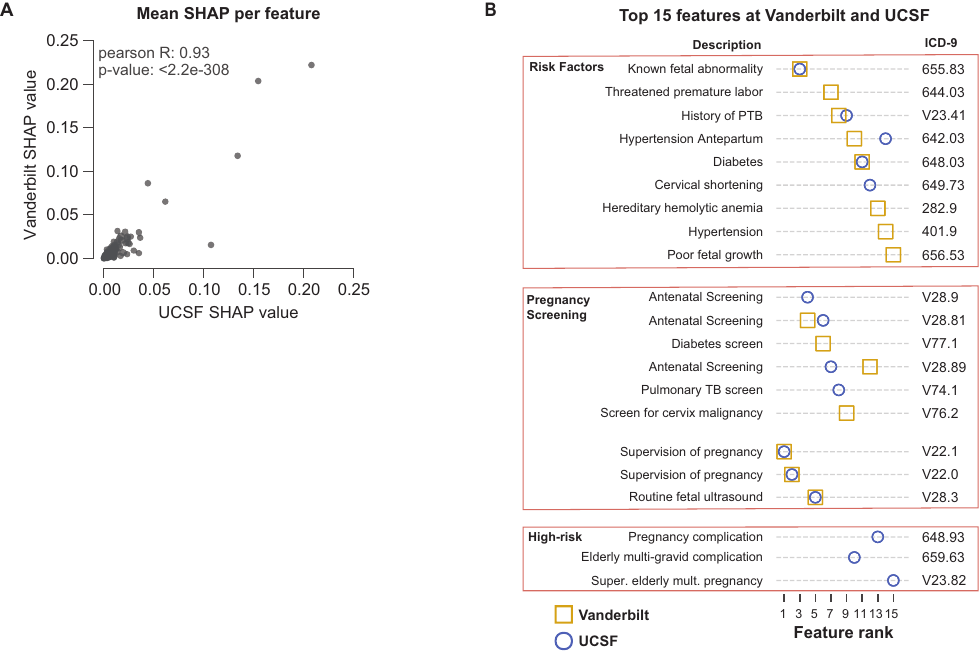

**Fig. S10: Preterm birth model feature importance is similar in an external cohort.** A preterm birth prediction model trained at Vanderbilt was applied to an external UCSF cohort. Models were trained using ICD-9 codes present before 28 weeks of gestation at Vanderbilt on 16,857 of women and evaluated on a held-out set at Vanderbilt (n=4,215, gold) and UCSF cohort (n=5,978, blue). These models performed similarly (Fig. 7). **(A)** Feature importance was estimated by the mean absolute SHapley Additive exPlanation (SHAP) value per feature in each individual in each cohort (x and y-axes). The feature importance estimates have a high positive correlation between cohorts (Pearson r=0.93, p<2.2e-308, two-tailed). **(B)** The top 15 features with the highest mean absolute SHAP score in the Vanderbilt cohort (gold square) or UCSF cohort (blue circle). The majority of the features were shared across cohorts and capture known risk factors (fetal abnormalities, history of preterm birth, etc.), pregnancy screening visits, and supervision of high-risk pregnancies.

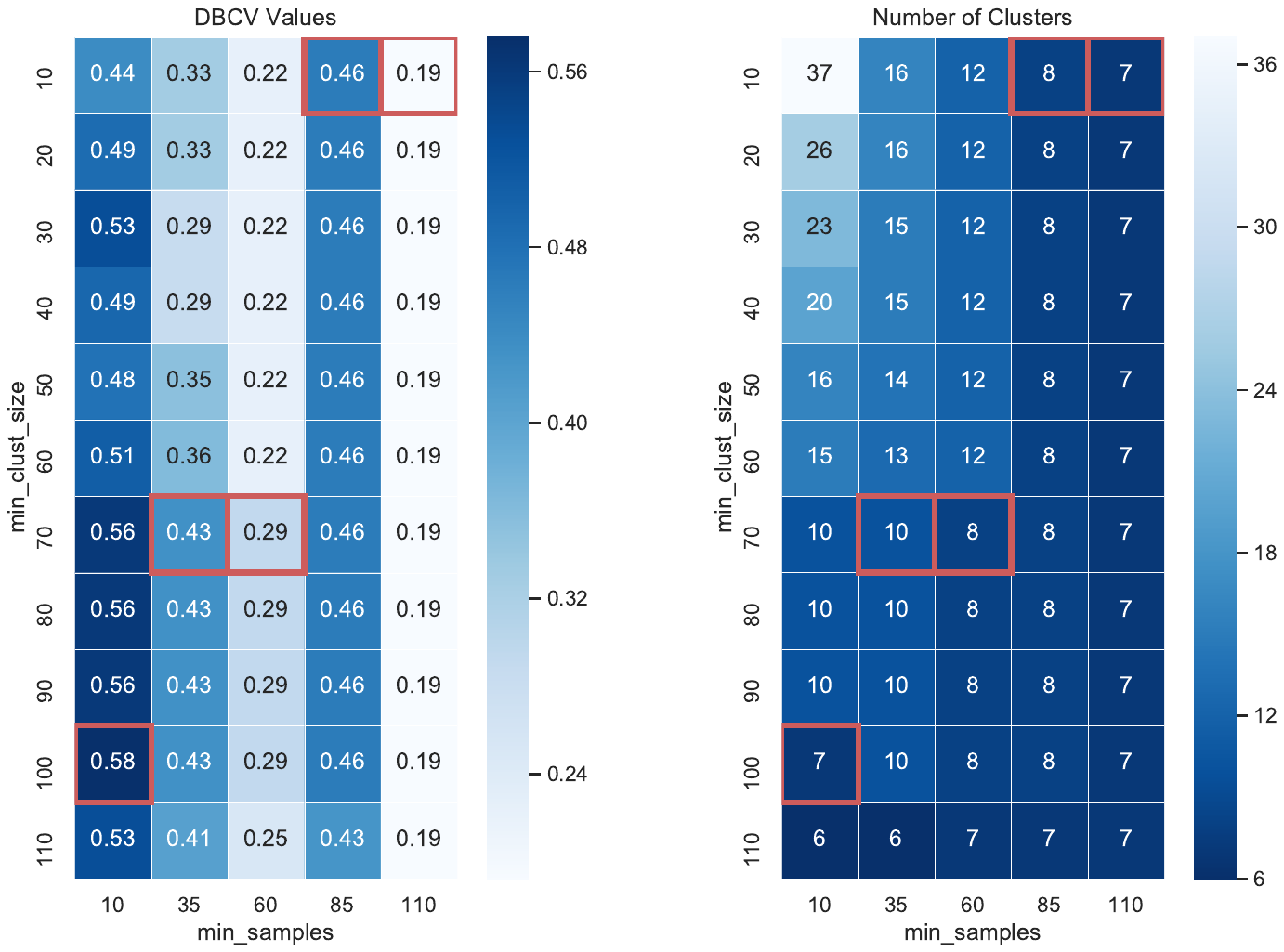

**Fig. S11: Density based cluster validity score across hyper-parameters space for HDBSCAN clustering of deliveries by feature importance.** To identify the optimum number of clusters using HDBSCAN on the billing-code-based model at 28 weeks gestation in the held-out set, we explored two hyperparameters: minimum number of individuals in each cluster (‘min_clust_size, y-axis) and threshold for determining outlier individuals who do not belong to a cluster (‘min_samples’, x-axis). The left heatmap represents cluster validity measured with the density-based cluster validity (DBCV) score with higher DBCV (darker blue) scores indicating more distinct clusters. The right heatmap displays the number of clusters (ligher blue == higher number of clusters) for the pair of hyperparameters. Cells outlined in red have the highest values within their column. Note, number of clusters includes a cluster for outliers.

**Supplemental Tables**

|  |  |  |  |  | **Number of Women** | | | |
| --- | --- | --- | --- | --- | --- | --- | --- | --- |
|  |  |  |  |  | **Race &** | **Not Race &** | **Race &** | **Not Race &** |
| **Race** | **Cluster** | **OR** | **log10(OR)** | **p-value** | **In Cluster** | **In Cluster** | **Not In Cluster** | **Not In Cluster** |
| WHITE | 1 | 0.853 | -0.069 | 0.5072080 | 68 | 34 | 1,503 | 641 |
| WHITE | 2 | 1.074 | 0.031 | 0.6356394 | 213 | 86 | 1,358 | 589 |
| WHITE | 3 | 1.064 | 0.027 | 0.8322583 | 79 | 32 | 1,492 | 643 |
| WHITE | 4 | 0.757 | -0.121 | 0.0972683 | 108 | 60 | 1,463 | 615 |
| WHITE | 5 | 1.242 | 0.094 | 0.0220511 | 673 | 254 | 898 | 421 |
| WHITE | 6 | 0.848 | -0.071 | 0.1111089 | 420 | 203 | 1,151 | 472 |
| AA | 1 | 1.246 | 0.096 | 0.3672195 | 23 | 79 | 406 | 1,738 |
| AA | 2 | 0.972 | -0.012 | 0.9370306 | 56 | 243 | 373 | 1,574 |
| AA | 3 | 1.246 | 0.095 | 0.3853035 | 25 | 86 | 404 | 1,731 |
| AA | 4 | 0.798 | -0.098 | 0.3581256 | 27 | 141 | 402 | 1,676 |
| AA | 5 | 0.976 | -0.011 | 0.8276443 | 175 | 752 | 254 | 1,065 |
| AA | 6 | 1.000 | 0.000 | 1.0000000 | 119 | 504 | 310 | 1,313 |
| ASIAN | 1 | 0.925 | -0.034 | 1.0000000 | 3 | 99 | 68 | 2,076 |
| ASIAN | 2 | 0.593 | -0.227 | 0.2861020 | 6 | 293 | 65 | 1,882 |
| ASIAN | 3 | 0.268 | -0.572 | 0.2594471 | 1 | 110 | 70 | 2,065 |
| ASIAN | 4 | 1.147 | 0.060 | 0.6497255 | 6 | 162 | 65 | 2,013 |
| ASIAN | 5 | 1.398 | 0.146 | 0.1783888 | 35 | 892 | 36 | 1,283 |
| ASIAN | 6 | 1.022 | 0.010 | 0.8937709 | 20 | 603 | 51 | 1,572 |
| HISPANIC | 1 | 1.008 | 0.003 | 1.0000000 | 8 | 94 | 167 | 1,977 |
| HISPANIC | 2 | 1.038 | 0.016 | 0.8176980 | 24 | 275 | 151 | 1,796 |
| HISPANIC | 3 | 0.665 | -0.177 | 0.4656009 | 6 | 105 | 169 | 1,966 |
| HISPANIC | 4 | 2.497 | 0.397 | 0.0002320 | 27 | 141 | 148 | 1,930 |
| HISPANIC | 5 | 0.452 | -0.345 | 0.0000044 | 44 | 883 | 131 | 1,188 |
| HISPANIC | 6 | 1.560 | 0.193 | 0.0082143 | 64 | 559 | 111 | 1,512 |

**Table S1: Enrichment of race per cluster in the held-out cohort for the model predicting preterm birth by 28 weeks of gestation.** We calculated enrichment (Odds Ratio, OR) for each cluster using Fisher’s exact test on the contingency table of number of women based on race (Race, Not Race) and cluster membership. The cohort was the held-out set used to evaluate the preterm birth prediction at 28 weeks of gestation.

|  |  |  |  |  | **Number of Women** | | | |
| --- | --- | --- | --- | --- | --- | --- | --- | --- |
|  |  |  |  |  | **High Risk &** | **Low Risk &** | **High Risk &** | **Low Risk &** |
| **Risk Factor** | **Cluster** | **OR** | **log10(OR)** | **p-value** | **In Cluster** | **In Cluster** | **Not In Cluster** | **Not In Cluster** |
| cerv_abnml | 1 | 2.022 | 0.306 | 2.81E-01 | 2 | 100 | 21 | 2,123 |
| cerv_abnml | 2 | 0.618 | -0.209 | 7.59E-01 | 2 | 297 | 21 | 1,926 |
| cerv_abnml | 3 | 0.873 | -0.059 | 1.00E+00 | 1 | 110 | 22 | 2,113 |
| cerv_abnml | 4 | 4.490 | 0.652 | 5.53E-03 | 6 | 162 | 17 | 2,061 |
| cerv_abnml | 5 | 0.499 | -0.302 | 2.00E-01 | 6 | 921 | 17 | 1,302 |
| cerv_abnml | 6 | 0.919 | -0.037 | 1.00E+00 | 6 | 617 | 17 | 1,606 |
| gest_dm | 1 | 2.065 | 0.315 | 7.56E-03 | 22 | 80 | 252 | 1,892 |
| gest_dm | 2 | 0.534 | -0.272 | 5.69E-03 | 22 | 277 | 252 | 1,695 |
| gest_dm | 3 | 9.330 | 0.970 | 2.25E-25 | 57 | 54 | 217 | 1,918 |
| gest_dm | 4 | 1.634 | 0.213 | 2.66E-02 | 30 | 138 | 244 | 1,834 |
| gest_dm | 5 | 0.727 | -0.138 | 1.84E-02 | 95 | 832 | 179 | 1,140 |
| gest_dm | 6 | 0.502 | -0.299 | 1.94E-05 | 47 | 576 | 227 | 1,396 |
| gest_htn | 1 | 1.686 | 0.227 | 1.60E-01 | 8 | 94 | 103 | 2,041 |
| gest_htn | 2 | 0.233 | -0.632 | 8.24E-04 | 4 | 295 | 107 | 1,840 |
| gest_htn | 3 | 26.425 | 1.422 | 9.06E-39 | 49 | 62 | 62 | 2,073 |
| gest_htn | 4 | 1.097 | 0.040 | 7.13E-01 | 9 | 159 | 102 | 1,976 |
| gest_htn | 5 | 0.419 | -0.378 | 6.66E-05 | 26 | 901 | 85 | 1,234 |
| gest_htn | 6 | 0.392 | -0.406 | 4.40E-04 | 15 | 608 | 96 | 1,527 |
| (pre)eclam | 1 | 2.245 | 0.351 | 1.26E-02 | 13 | 89 | 131 | 2,013 |
| (pre)eclam | 2 | 0.989 | -0.005 | 1.00E+00 | 19 | 280 | 125 | 1,822 |
| (pre)eclam | 3 | 3.564 | 0.552 | 1.32E-05 | 20 | 91 | 124 | 2,011 |
| (pre)eclam | 4 | 0.918 | -0.037 | 1.00E+00 | 10 | 158 | 134 | 1,944 |
| (pre)eclam | 5 | 0.957 | -0.019 | 8.61E-01 | 58 | 869 | 86 | 1,233 |
| (pre)eclam | 6 | 0.476 | -0.323 | 9.91E-04 | 23 | 600 | 121 | 1,502 |
| sickle_cell | 1 | 0.000 |  | 1.00E+00 | - | 102 | 20 | 2,124 |
| sickle_cell | 2 | 0.341 | -0.468 | 5.04E-01 | 1 | 298 | 19 | 1,928 |
| sickle_cell | 3 | 2.158 | 0.334 | 2.60E-01 | 2 | 109 | 18 | 2,117 |
| sickle_cell | 4 | 2.204 | 0.343 | 1.84E-01 | 3 | 165 | 17 | 2,061 |
| sickle_cell | 5 | 0.607 | -0.217 | 3.66E-01 | 6 | 921 | 14 | 1,305 |
| sickle_cell | 6 | 1.746 | 0.242 | 2.17E-01 | 8 | 615 | 12 | 1,611 |
| delivery_age | 1 | 1.832 | 0.263 | 1.09E-02 | 28 | 74 | 367 | 1,777 |
| delivery_age | 2 | 1.238 | 0.093 | 1.66E-01 | 61 | 238 | 334 | 1,613 |
| delivery_age | 3 | 1.796 | 0.254 | 1.03E-02 | 30 | 81 | 365 | 1,770 |
| delivery_age | 4 | 2.197 | 0.342 | 2.11E-05 | 51 | 117 | 344 | 1,734 |
| delivery_age | 5 | 0.694 | -0.158 | 1.61E-03 | 135 | 792 | 260 | 1,059 |
| delivery_age | 6 | 0.693 | -0.159 | 5.29E-03 | 87 | 536 | 308 | 1,315 |
| diabetes | 1 | 1.494 | 0.174 | 3.56E-01 | 4 | 98 | 57 | 2,087 |
| diabetes | 2 | 0.215 | -0.667 | 1.28E-02 | 2 | 297 | 59 | 1,888 |
| diabetes | 3 | 81.819 | 1.913 | 8.17E-48 | 44 | 67 | 17 | 2,118 |
| diabetes | 4 | 1.108 | 0.044 | 8.03E-01 | 5 | 163 | 56 | 2,022 |
| diabetes | 5 | 0.096 | -1.018 | 1.04E-09 | 4 | 923 | 57 | 1,262 |
| diabetes | 6 | 0.085 | -1.069 | 7.75E-07 | 2 | 621 | 59 | 1,564 |
| fetal_abnl | 1 | 8.535 | 0.931 | 2.07E-10 | 19 | 83 | 56 | 2,088 |
| fetal_abnl | 2 | 0.662 | -0.179 | 3.87E-01 | 7 | 292 | 68 | 1,879 |
| fetal_abnl | 3 | 2.046 | 0.311 | 9.47E-02 | 7 | 104 | 68 | 2,067 |
| fetal_abnl | 4 | 2.205 | 0.343 | 2.41E-02 | 11 | 157 | 64 | 2,014 |
| fetal_abnl | 5 | 0.375 | -0.426 | 2.96E-04 | 16 | 911 | 59 | 1,260 |
| fetal_abnl | 6 | 0.537 | -0.270 | 4.82E-02 | 13 | 610 | 62 | 1,561 |
| prepreg_bp | 1 | 1.304 | 0.115 | 2.23E-01 | 54 | 48 | 993 | 1,151 |
| prepreg_bp | 2 | 0.714 | -0.146 | 8.85E-03 | 118 | 181 | 929 | 1,018 |
| prepreg_bp | 3 | 2.720 | 0.435 | 7.86E-07 | 77 | 34 | 970 | 1,165 |
| prepreg_bp | 4 | 1.072 | 0.030 | 6.88E-01 | 81 | 87 | 966 | 1,112 |
| prepreg_bp | 5 | 0.771 | -0.113 | 2.64E-03 | 397 | 530 | 650 | 669 |
| prepreg_bp | 6 | 1.256 | 0.099 | 1.60E-02 | 316 | 307 | 731 | 892 |
| prepreg_bmi | 1 | 1.203 | 0.080 | 3.63E-01 | 52 | 50 | 994 | 1,150 |
| prepreg_bmi | 2 | 0.541 | -0.267 | 2.00E-06 | 101 | 198 | 945 | 1,002 |
| prepreg_bmi | 3 | 2.850 | 0.455 | 2.65E-07 | 78 | 33 | 968 | 1,167 |
| prepreg_bmi | 4 | 1.390 | 0.143 | 4.43E-02 | 91 | 77 | 955 | 1,123 |
| prepreg_bmi | 5 | 0.762 | -0.118 | 1.71E-03 | 395 | 532 | 651 | 668 |
| prepreg_bmi | 6 | 1.317 | 0.119 | 3.94E-03 | 321 | 302 | 725 | 898 |

**Table S2: Enrichment of clinical risk factors per cluster in the held-out cohort for the model predicting preterm birth by 28 weeks of gestation.** We calculated enrichment (Odds Ratio, OR) for each cluster using Fisher’s exact test on the contingency table of the number of women based on clinical risk factor status (High-Risk, Low-Risk) and cluster membership (Methods). The cohort was the held-out set used to evaluate the model predicting preterm birth at 28 weeks of gestation.

|  | **UCSF** | | | | **Vanderbilt** | | |
| --- | --- | --- | --- | --- | --- | --- | --- |
|  | **Not-Preterm** | **Preterm** | **p-value** |  | **Not-Preterm** | **Preterm** | **p-value** |
| **n** | 5615 | 363 |  |  | 18,498 | 2,651 |  |
| **Patient Age (mean (SD))** | 36.65 (5.08) | 36.54 (5.96) | 0.691 |  | 27.71 (5.75) | 27.73 (6.38) | 0.876 |
| **Patient Race (%)** |  |  | <0.001 |  |  |  | <0.001 |
| **American Indian or Alaska Native** | 26 (0.5) | 3 (0.8) |  |  | 47 (0.2) | 4 (0.01) |  |
| **Asian** | 1,336 (23.8) | 51 (14.0) |  |  | 1,051 (5.8) | 100 (3.8) |  |
| **Black or African American** | 336 (6.0) | 31 (8.5) |  |  | 2,962 (16.5) | 486 (18.8) |  |
| **Declined** | 72 (1.3) | 5 (1.4) |  |  | NA | NA |  |
| **Native Hawaiian/Pacific Islander** | 86 (1.5) | 3 (0.8) |  |  | NA | NA |  |
| **Other** | 866 (15.4) | 77 (21.2) |  |  | 162 (0.9) | 12 (0.04) |  |
| **Unknown** | 200 (3.6) | 32 (8.8) |  |  | 619 (3.3) | 69 (2.6) |  |
| **White or Caucasian** | 2,693 (48.0) | 161 (44.4) |  |  | 11,278(63.0) | 1,658 (64.2) |  |

**Table S3: Demographic distribution of UCSF and Vanderbilt cohorts.** We identified women with preterm and not preterm deliveries at UCSF and Vanderiblt using similar ascertainment (Methods). For each woman, we predicted the earliest delivery in their EHR. We report age at delivery (Patient Age) and self- or third-party reported race for both cohorts. T-tests and chi-squared tests of independence were used to compare distrbutions stratfied by delivery label.

| **icd9** | **shap** | **icd9** | **shap** | **icd9** | **shap** | **icd9** | **shap** | **icd9** | **shap** | **icd9** | **shap** |
| --- | --- | --- | --- | --- | --- | --- | --- | --- | --- | --- | --- |
| V22.1 | 2.2E-01 | 311 | 6.4E-03 | 780.79 | 2.8E-03 | 367.1 | 1.5E-03 | 959.4 | 8.8E-04 | 719.46 | 4.9E-04 |
| V22.0 | 2.0E-01 | 216.5 | 6.4E-03 | 786.2 | 2.8E-03 | 620.1 | 1.5E-03 | 795.05 | 8.6E-04 | 780.4 | 4.8E-04 |
| 655.83 | 1.2E-01 | V74.1 | 6.2E-03 | 706.1 | 2.8E-03 | 789.06 | 1.5E-03 | 304 | 8.6E-04 | V15.81 | 4.8E-04 |
| V28.81 | 8.6E-02 | 626.4 | 6.1E-03 | V15.89 | 2.8E-03 | V25.01 | 1.4E-03 | 883 | 8.6E-04 | 714 | 4.5E-04 |
| V28.3 | 6.5E-02 | 641.03 | 6.1E-03 | 462 | 2.8E-03 | 704.8 | 1.4E-03 | 626.1 | 8.6E-04 | 659.71 | 4.5E-04 |
| V77.1 | 3.1E-02 | 276.51 | 6.1E-03 | 641.01 | 2.8E-03 | V06.8 | 1.4E-03 | V72.2 | 8.4E-04 | 656.23 | 4.1E-04 |
| 644.03 | 3.1E-02 | 626 | 5.9E-03 | 486 | 2.6E-03 | 789.59 | 1.4E-03 | 728.85 | 8.4E-04 | V25.8 | 4.1E-04 |
| V23.41 | 3.0E-02 | V72.42 | 5.9E-03 | V67.09 | 2.6E-03 | 789.04 | 1.4E-03 | 628.9 | 8.2E-04 | 789.05 | 3.9E-04 |
| V76.2 | 2.6E-02 | 789.09 | 5.3E-03 | 309.81 | 2.6E-03 | 655.91 | 1.4E-03 | 709.09 | 8.2E-04 | 788.41 | 3.7E-04 |
| 642.03 | 2.5E-02 | V72.40 | 5.3E-03 | 789.01 | 2.6E-03 | 794.5 | 1.4E-03 | 34 | 8.1E-04 | 790.6 | 3.3E-04 |
| 648.03 | 2.5E-02 | V23.82 | 5.0E-03 | 616.1 | 2.5E-03 | 655.93 | 1.4E-03 | 309.28 | 8.0E-04 | 304.01 | 3.2E-04 |
| V28.89 | 2.4E-02 | 787.01 | 5.0E-03 | 656.73 | 2.5E-03 | 564 | 1.3E-03 | 466 | 7.9E-04 | V71.89 | 2.9E-04 |
| 282.9 | 2.3E-02 | 623.5 | 5.0E-03 | 256.4 | 2.4E-03 | 789.03 | 1.3E-03 | 520.6 | 7.9E-04 | 780.39 | 2.6E-04 |
| 401.9 | 2.3E-02 | 795.01 | 4.9E-03 | 658.13 | 2.3E-03 | 783.21 | 1.3E-03 | 276.2 | 7.9E-04 | 786.59 | 2.2E-04 |
| 656.53 | 2.1E-02 | 278 | 4.7E-03 | 724.2 | 2.3E-03 | 296.9 | 1.3E-03 | 719.41 | 7.4E-04 | 648.01 | 2.0E-04 |
| V72.31 | 1.9E-02 | 648.81 | 4.6E-03 | 641.13 | 2.3E-03 | 648.33 | 1.3E-03 | 654.53 | 7.2E-04 | 478 | 1.7E-04 |
| 642.23 | 1.9E-02 | 465.9 | 4.5E-03 | 648.31 | 2.3E-03 | 977.9 | 1.3E-03 | 729.5 | 7.2E-04 | 250.02 | 1.4E-04 |
| 648.93 | 1.9E-02 | 787.02 | 4.2E-03 | 659.61 | 2.2E-03 | E917.9 | 1.2E-03 | 626.8 | 7.1E-04 | V23.7 | 9.8E-05 |
| V23.89 | 1.8E-02 | V70.0 | 4.2E-03 | 719.45 | 2.2E-03 | 642.01 | 1.2E-03 | 655.01 | 7.0E-04 |  |  |
| 649.73 | 1.7E-02 | V25.09 | 4.1E-03 | 787.91 | 2.1E-03 | V26.49 | 1.2E-03 | 595 | 6.9E-04 |  |  |
| 625.9 | 1.6E-02 | 790.29 | 4.0E-03 | 634.92 | 2.1E-03 | 786.05 | 1.2E-03 | 648 | 6.9E-04 |  |  |
| 649.13 | 1.6E-02 | 655.13 | 3.8E-03 | 719.47 | 2.1E-03 | 461.9 | 1.2E-03 | 610.1 | 6.9E-04 |  |  |
| V28.9 | 1.5E-02 | 244.9 | 3.8E-03 | 655.73 | 2.1E-03 | V67.00 | 1.2E-03 | 719.44 | 6.7E-04 |  |  |
| 250.01 | 1.5E-02 | 780.2 | 3.8E-03 | V45.89 | 2.1E-03 | 782.3 | 1.2E-03 | 648.91 | 6.7E-04 |  |  |
| 640.83 | 1.4E-02 | 477 | 3.8E-03 | 79.99 | 2.1E-03 | 813.42 | 1.1E-03 | 796.5 | 6.7E-04 |  |  |
| 655.03 | 1.3E-02 | 642.91 | 3.7E-03 | 737.3 | 2.1E-03 | V58.69 | 1.1E-03 | 790.22 | 6.7E-04 |  |  |
| 648.83 | 1.3E-02 | 250.03 | 3.7E-03 | 632 | 2.0E-03 | 305.1 | 1.1E-03 | 648.23 | 6.7E-04 |  |  |
| 654.23 | 1.2E-02 | 796.2 | 3.7E-03 | 285.9 | 1.9E-03 | 54.9 | 1.1E-03 | 729.81 | 6.6E-04 |  |  |
| 640.03 | 1.2E-02 | 640.93 | 3.6E-03 | 626.9 | 1.9E-03 | V05.9 | 1.1E-03 | 238.2 | 6.5E-04 |  |  |
| V27.0 | 1.2E-02 | 382.9 | 3.6E-03 | 620.2 | 1.9E-03 | 427.89 | 1.1E-03 | V70.7 | 6.4E-04 |  |  |
| 692.9 | 1.1E-02 | 655.81 | 3.6E-03 | 648.43 | 1.9E-03 | 611.72 | 1.1E-03 | 643.03 | 6.3E-04 |  |  |
| 782.1 | 1.1E-02 | V20.2 | 3.5E-03 | 795.03 | 1.9E-03 | V58.67 | 1.1E-03 | 305 | 6.3E-04 |  |  |
| 623.8 | 1.1E-02 | 616 | 3.5E-03 | 724.5 | 1.8E-03 | V72.83 | 1.1E-03 | 521 | 6.3E-04 |  |  |
| V74.5 | 1.0E-02 | 649.03 | 3.4E-03 | 845 | 1.8E-03 | 530.81 | 1.1E-03 | 478.19 | 6.1E-04 |  |  |
| 658.03 | 9.5E-03 | 788.1 | 3.4E-03 | 493.9 | 1.8E-03 | 473.9 | 1.0E-03 | 782 | 6.1E-04 |  |  |
| 659.63 | 9.2E-03 | V23.9 | 3.4E-03 | 641.93 | 1.7E-03 | 558.9 | 1.0E-03 | 218.9 | 5.9E-04 |  |  |
| V23.5 | 8.7E-03 | 658.01 | 3.4E-03 | 959.7 | 1.7E-03 | 780.5 | 1.0E-03 | 296.2 | 5.8E-04 |  |  |
| V22.2 | 8.6E-03 | 784 | 3.3E-03 | 620.8 | 1.7E-03 | 706.2 | 1.0E-03 | 698.1 | 5.8E-04 |  |  |
| 646.83 | 8.3E-03 | 795.07 | 3.2E-03 | 477.8 | 1.7E-03 | 659.53 | 1.0E-03 | 280 | 5.6E-04 |  |  |
| V04.81 | 8.3E-03 | 642.93 | 3.2E-03 | 646.63 | 1.6E-03 | 646.81 | 1.0E-03 | 401.1 | 5.4E-04 |  |  |
| 789 | 8.3E-03 | 786.5 | 3.2E-03 | 346.9 | 1.6E-03 | 924.9 | 1.0E-03 | 611.71 | 5.3E-04 |  |  |
| 278.01 | 7.9E-03 | 286.9 | 3.2E-03 | 656.51 | 1.6E-03 | V25.49 | 9.8E-04 | 622.11 | 5.3E-04 |  |  |
| V25.42 | 7.9E-03 | 649.53 | 3.2E-03 | V15.85 | 1.6E-03 | 218.1 | 9.8E-04 | 296.31 | 5.3E-04 |  |  |
| 599 | 7.8E-03 | 112.1 | 3.1E-03 | 477.9 | 1.6E-03 | 625 | 9.6E-04 | V72.84 | 5.2E-04 |  |  |
| V23.49 | 7.2E-03 | 649.11 | 3.0E-03 | 659.73 | 1.6E-03 | V65.49 | 9.5E-04 | 644.1 | 5.2E-04 |  |  |
| 250 | 7.0E-03 | 649.63 | 3.0E-03 | 646.23 | 1.5E-03 | 795.04 | 9.4E-04 | 300.01 | 5.2E-04 |  |  |
| 654.43 | 6.7E-03 | V28.4 | 2.9E-03 | 276.8 | 1.5E-03 | 785 | 9.0E-04 | 611.79 | 5.1E-04 |  |  |
| V72.9 | 6.5E-03 | 780.6 | 2.9E-03 | 787.03 | 1.5E-03 | 656.63 | 8.9E-04 | 786.09 | 5.0E-04 |  |  |
| 654.13 | 6.4E-03 | 648.13 | 2.8E-03 | 648.63 | 1.5E-03 | V77.91 | 8.9E-04 | 626.2 | 5.0E-04 |  |  |

**Table S4: All features (ICD-9 codes) used to predict preterm birth at 28 weeks using Vanderbilt cohort.** We report the mean absoulte SHAP value (‘shap’) across women in the held-out set for ICD-9 codes (‘icd9’) used to predict preterm birth at 28 weeks of gestation in the Vanderbilt cohort. Features with zero mean absolute SHAP value are not reported in the table.

**Supplementary Materials and Methods**

*Delivery-specific ICD-9/10 codes used to ascertain delivery type.*

The following ICD-9/10 codes were used to ascertain delivery type as described in the Methods section.

- Preterm Birth: 'O60.1 ', 'O60.10', 'O60.10X0', 'O60.10X1', 'O60.10X2', 'O60.10X3', 'O60.10X4', 'O60.10X5', 'O60.10X9', 'O60.12', 'O60.12X0', 'O60.12X1', 'O60.12X2', 'O60.12X3', 'O60.12X4', 'O60.12X5', 'O60.12X9', 'O60.13', 'O60.13X0', 'O60.13X1', 'O60.13X2', 'O60.13X3', 'O60.13X4', 'O60.13X5', 'O60.13X9', 'O60.14', 'O60.14X0', 'O60.14X1', 'O60.14X2', 'O60.14X3', 'O60.14X4', 'O60.14X5', 'O60.14X9', '644.2', '644.20', '644.21'
- Term Birth: 'O60.20', 'O60.20X0', 'O60.20X1', 'O60.20X2', 'O60.20X3', 'O60.20X4', 'O60.20X5', 'O60.20X9', 'O60.22', 'O60.22X0', 'O60.22X1', 'O60.22X2', 'O60.22X3', 'O60.22X4', 'O60.22X5', 'O60.22X9', 'O60.23', 'O60.23X0', 'O60.23X1', 'O60.23X2', 'O60.23X3', 'O60.23X4', 'O60.23X5', 'O60.23X9', 'O80', 'O48.0', '650', '645.1', '645.10', '645.11', '645.13', '649.8', '649.81', '649.82'
- Postterm Birth: 'O48.1', '645.2', '645.20', '645.21', '645.23', '645.00', '645.01', '645.03'

*Delivery-specific CPT codes used to ascertain delivery type.*

The following CPT codes were used to ascertain delivery date: '59400', '59409', '59410', '59414', '59510', '59514', '59515', '59525', '59610', '59612', '59614', '59618', '59620', '59622'.

*Identifying multiple gestations using billing codes.*

Pregnancies with multiple gestations were identified using the presence of any of the following billing codes. For singleton only analyses, we excluded women with multiple gestation.

- ICD-9 Multiple Gestations: '651','651.7','651.70','651.71','651.8','651.81','651.83','651.9','651.91','651.93','652.6','652.60','652.61','652.63','V91','V91.9','V91.90','V91.91','V91.92','V91.99',’651’,’651.0’,’651.00’,’651.01’,’651.03’,’651.1’,’651.10’,’651.11’,’651.13’,’651.2’,’651.20’,’651.21’,’651.23’,’651.3’,’651.30’,’651.31’,’651.33’,’651.4’,’651.40’,’651.41’,’651.43’,’651.5’,’651.50’, ‘651.51’, ‘651.53’, ‘V91’,’V91.0’,’V91.00’,’V91.01’,’V91.02’,’V91.03’,’V91.09’,’V91.1’,’V91.10’,’V91.11’,’V91.12’,’V91.19’,’V91.2’,’V91.20’,’V91.21’,’V91.22’,’V91.29’,’V91.9’,’V91.90’,’V91.91’,’V91.92’, ‘V91.99’
- CPT Twin codes: ‘74713’,’76802’,’76810’,’76812’,’76814’
- ICD-10 Multiple Gestations: 'BY4BZZZ','BY4DZZZ','BY4GZZZ','O30.801','O30.802','O30.803','O30.809','O30.811','O30.812','O30.813','O30.819','O30.821','O30.822','O30.823','O30.829','O30.891','O30.892','O30.893','O30.899','O30.91','O30.92','O30.93','O31.BX10','O31.BX11','O31.BX12','O31.BX13','O31.BX14','O31.BX15','O31.BX19','O31.BX20','O31.BX21','O31.BX22','O31.BX23','O31.BX24','O31.BX25','O31.BX29','O31.BX30','O31.BX31','O31.BX32','O31.BX33','O31.BX34','O31.BX35','O31.BX39','O31.BX90','O31.BX91','O31.BX92','O31.BX93','O31.BX94','O31.BX95','O31.BX99'

*Past medical and family history extracted from EHRs used to predict preterm birth.*

The following past medical and family history features were extracted from EHRs for women with at least one recorded delivery at Vanderbilt Hospital.

- Maternal History:

'Abortion', 'Alcohol ', 'Baby's father had a child with birth defect not listed', 'Baby's father's family has history of birth defect not listed', 'Drugs ', 'Endocrine Metabolic Patient ', 'Endocrine metaboloic Patient History ', 'Gravidity', 'Hematoligic ', 'Maternal metabolic or endocrine disorders (Diabetes, PKU) ', 'Menses every 28 to 30 days ', 'Patient History Breast Disease ', 'Patient History Congential Heart Defect ', 'Patient History Cystic Fibrosis ', 'Patient History Down Syndrome ', 'Patient History GI Problems ', 'Patient History Genetic other', 'Patient History Gyn Problems ', 'Patient History Heart Disease ', 'Patient History Hemophilia or other blood disorders ', 'Patient History Huntington's Chorea ', 'Patient History Hypertension ', 'Patient History Immune or Infectious Disease ', 'Patient History Infertility or Recurrent Spontaneous Abortions ', 'Patient History Malignancies ', 'Patient History Mental Retardation ', 'Patient History Multiple births ', 'Patient History Muscular Dystrophy ', 'Patient History Neural Tube Defect ', 'Patient History Neurological Disorder ', 'Patient History Operations or Accidents ', 'Patient History Other Hospitalizations ', 'Patient History Other ', 'Patient History Other inherited or chromosomal disorders ', 'Patient History Other structural Birth defect ', 'Patient History Phlebitis or varicocities ', 'Patient History Pulmonary Disease ', 'Patient History Recurrent Pregnancy loss defined as more than 2 or stillbirth', 'Patient History STDs ', 'Patient History Sickle Cell Disease (African or Carribean American) ', 'Patient History Thalessemia (Italian, Greek, Mediterranean, or Asian Background); MCV <80 ', 'Patient History Tobacco, Alcohol, Drugs ', 'Patient History Urinary tract problems including UTIs and Pyel ', 'Patient History of Seizure', 'Patient History of sexual/physical abuse or trauma ', 'Patient's age greater than 34 at delivery ', 'Pregnancy Induced Hypertension', 'Prior Preterm_births', 'Regular exercise ', 'Term_births', 'Tobacco ', 'Urinary tract infection', 'Live_Children'

- Family History:

'Familly History Thalessemia (Italian, Greek, Mediterranean, or Asian Background); MCV <80 ', 'Family History Breast Disease ', 'Family History Congential Heart Defect ', 'Family History Cystic Fibrosis ', 'Family History Down Syndrome ', 'Family History GI Problems ', 'Family History Genetic other', 'Family History Gyn Problems ', 'Family History Heart Disease ', 'Family History Hemophilia or other blood disorders ', 'Family History Huntington's Chorea ', 'Family History Hypertension ', 'Family History Immune or Infectious Disease ', 'Family History Infertility or Recurrent Spontaneous Abortions ', 'Family History Jewish, Cajun, French Canadian (Tay Sachs) ', 'Family History Jewish: Canavan Disease, Gauchers ', 'Family History Malignancies ', 'Family History Mental Retardation ', 'Family History Metabolic or endocrine disorders (Diabetes, PKU) ', 'Family History Multiple births ', 'Family History Muscular Dystrophy ', 'Family History Neural Tube Defect ', 'Family History Neuroligcal Disorder ', 'Family History Operations or Accidents ', 'Family History Other Hospitalizations ', 'Family History Other ', 'Family History Other inherited or chromosomal disorders ', 'Family History Other structural Birth defect ', 'Family History Phlebitis or varicocities ', 'Family History Pulmonary Disease ', 'Family History Recurrent Pregnancy loss defined as more than 2 or stillbirth', 'Family History STDs ', 'Family History Sickle Cell Disease (African or Carribean American) ', 'Family History Tobacco, Alcohol, Drugs ', 'Family History Urinary tract problems including UTIs and Pyel ', 'Family History of Seizure', 'Family History of sexual/physical abuse or trauma ', 'Jewish, Cajun, French Canadian (Tay Sachs) ', 'Jewish: Canavan Disease, Gauchers'

*Clinical labs measured during pregnancy used to predict preterm birth*

'albumin urine, lactic acid venous, cd3 #/cumm, total protein urine, glucose blood, wbc blood, eo automated abs, atyp lymphs (abs), reaction time, lmw heparin assay, rdwsd, glucose spinal fluid, control ptt, rbc folate, calcium blood, gentamicin level, urea nitrogen ur spot, mch, aldosterone, magnesium blood, mchc, factor viii activity, sodium blood, igg quantitative blood, bicarbonate (calc), hcg beta (3rd irp), dhea sulfate, hdl cholesterol, protein csf, f t4, alt blood, neutrophil %, k-time, metamyelo %, estriol unconjugated, sodium urine spot, cellano antigen, icterus index, nucleated rbc, protein total blood, eosoinophil (abs), erythropoietin, neutrophils %, immature retic fraction, zinc serum, c-peptide, imm granulocytes %, lipemia index, monocytes %, ssb (la)(ena) ab, igg, beta-hcg serum, protein urine, bedside glucose, troponin t, intact-pth, sm (smith) autoabs eia, ferritin, absolute cd8, sex hormone bind globulin, eosinophils %, protein c activity, cd8(cd3+)/cd45 #/cumm, glucose tol 50g, basophils %, wbc, albumin, mcv, gamma globulin, testosterone free, fio2, lymph %, pan t cd3 %, troponin-i, mono (abs), rheumatoid factor, quant d-dimer for dic, pcv blood, hgb a1c glycated poc, 25-hydroxy d3, eosoinophil (abs), carboxyhemoglobin, urea nitrogen blood, hgb a1c glycated, cholesterol blood, lamotrigine, cystatin-c, carbon dioxide blood, apa-igg, neutrophils %, myelocytes %, hdl cholesterol, vit e(alpha-tocopherol), glucose whole blood, calcium ionized, gamma glut trans blood, follicle stimulating hrm, total hemoglbin, creatinine g/24 hour, atyp lymphs %, wbc urine micro, nt automated abs, chloride blood, imm platelet fraction, fasting glucose, po2/fio2, sodium whole blood, ast blood, albumin/creatinine ratio, angle(alpha), rbc, vit d 1,25-dihydroxy, c3 quantitative blood, lymphs (abs), ldl cholesterol, triglycerides blood, testosterone, ed troponin-i wbld, o2 saturation, creatinine urine per day, triiodothyronine free, eosinophil %, rbc, rbc urine micro, thyroid stim hormone, anti-myeloperoxidase, c-reactive protein, deamidated gliadin iga abs, hyaline cast, ammonia, igg beta 2 glycoprotein i, progesterone rapid, vitamin d 25-oh total, t helper cd4 #/cumm, patient (pt), schedule q hr, keppra (levetiracetam), creatine kinase total, maternal alphafeto pr0, creatinine urine "spot", ret ct, creatinine urine "timed", specific gravity ua, iron blood, kappa light chain quant, lithium blood, 2 hour glucose, vancomycin level, anion gap, luteinizing hormone, iga quantitative blood, phenytoin (dilantin), methemoglobin, alpha-1 globulin, thyroglobulin serum, renin activity, c4 quantitative blood, rdw, urobilinogen, maternal weight, venous ph, % cd3, protein urine spot, carbamazepine (tegretol), hep b surface ab value, anti-protease 3, hemoglobin s, sed rate, amylase blood, ssa (ro)(ena) ab, igg, 25-hydroxy d total, total gamma globulin, adrenocorticotropic horm, retic hgb equiv, neut (abs), insulin, albumin, lymphs %, antithrombin iii act, myelocytes (abs), lymps %, nucleated rbc, alkaline phosphatase bld, # wbc\'s counted, fibrinogen, ed creatinine wbld, ph arterial, metamyelocytes (abs), kappa/lambda ratio, ret abs, beta globulin, basophils %, albumin blood, ed inr wbld, anti thyroid peroxab, tc:hdl ratio, afp tumor, vitamin a (retinol), albumin/creat ratio, patient location, ck-mb ratio, total volume, total t4, creatinine blood, absolute cd3, collection time, current gest age, apa-igm, ck blood, hemoglobin blood, max amplitude, transferrin blood, cd8(cd3+)/cd45 %, cd4:cd8 ratio, monocytes %, protein urine timed, beta globulin, dose, % cd8, estradiol, nucleated rbc#, cortisol, prolactin, lymphs (abs), granular cast, protein-s-activity, pcv blood, mono (abs), brain natriuretic peptide, fk-506 (tacrolimus), bilirubin conjugated, bilirubin total blood, chloride whole blood, 25-hydroxy d2, hemoglobin a, haptoglobin blood, folate serum, ck-mb, glucose urine, nucleated cell, absolute cd4, baso (abs), creatinine urine, scl-70 autoabs eia, infusion start time, squamous epithelial, g parameter, osmolality blood, baso (abs), vitamin b-12, hours of collection, inr, lipase blood, hemoglobin a2, potassium urine spot, factor v leiden coag, phosphorus inorganic, percent saturation, valproate(depakane), ldh blood, anti-dna(sle)current, lambda light chain quant, bcrabl/bcr ratio, free phenytoin, t helper cd4 %, % cd4, mean platelet volume, creatinine urine, ketone urine, igm quantitative blood, patient ptt, glucose body fluid, vit e(gamma-tocopherol), igm beta 2 glycoprotein i, maternal b-hcg, drvvt, protein total blood, ph urine, lymph abs, alpha-2 globulin, retinyl palminate, d-dimer (patient), lymphs %, o2 saturation (calc), uric acid blood, ldl cholesterol calc, non-hdl, triiodothyronine, total, testerone free female, potassium whole blood, deamidated gliadin igg abs, urobilinogen, monocytes %, protein /24 hour, neut (abs), tibc blood, apa-iga, platelet count, albumin urine, o2 saturation(venous, prealbumin blood, basophils %, eosinophil %

*Identifying cesarean section and vaginal deliveries.*

The following ICD-9 and CPT codes were used to label deliveries as a cesarean section vs. vaginal deliveries. We excluded deliveries if they had codes for both types of deliveries.

- Cesarean section: '669.7', '669.70', '669.71', '763.4', '74.0', '74.1', '74.2', '74.4', '74.9','74.99', '59510', '59514', '59515', '59618', '59620', '59622'
- Vaginal Deliveries: '59409', '59410', '59610', '59612', '59614'

*Identifying spontaneous preterm births from electronic health records*

From all preterm cases, we excluded women meeting any of the following criteria: medically induced labor, delivery by cesarean section, or preterm premature rupture of membranes. The following ICD-9 and CPT codes were used to identify these exclusion criteria.

- Medically induced labor: '73.01', '73.1', '73.4', '73.0', '73.09'
- Cesarean Section delivery: '669.7', '669.70', '669.71', '763.4', '74.0', '74.1', '74.2', '74.4', '74.9','74.99', '59510', '59514', '59515', '59618', '59620', '59622'
- Preterm premature rupture of membranes: '658.13','658.10','658.11'

*Identifying clinical risk factors based on ICD-9 codes*

Women with at least one ICD-9 code within each risk factor set was considered to be positive for that risk factor.

Fetal abnormalities: '655','655.01','655.03','655.1','655.1','655.11','655.13','655.3','655.31','655.33','655.4','655.41','655.43','655.5','655.5','655.51','655.53'

Sickle Cell Disease:

'282.41','282.42','282.5','282.6','282.6','282.61','282.62','282.63','282.64','282.68','282.69'

Diabetes Codes:

'250','250.01','250.02','250.03','250.1','250.11','250.12','250.13','250.2','250.21','250.22','250.23','250.3','250.31','250.32','250.33','250.4','250.41','250.42','250.43','250.5','250.51','250.52','250.53','250.6','250.61','250.62','250.63','250.7','250.71','250.72','250.73','250.8','250.81','250.82','250.83','250.9','250.91','250.92','250.93'

Gestational Diabetes Codes:

"648.0","648.00","649.1","648.01","649.1","648.02","649.1","648.03","649.1","648.04"

Gestational Hypertension Codes:

"642","642.0","642.00","642.01","642.02","642.03","642.04","642.1","642.10","642.11","642.12","642.13","642.14","642.2","642.20","642.21","642.22","642.23","642.24","642.3","642.30","642.31","642.32","642.33","642.34"

Eclampsia or Preeclampsia Codes:

"642.4","642.40","642.41","642.42","642.43","642.44","642.5","642.50","642.51","642.52","642.53","642.54","642.6","642.60","642.61","642.62","642.63","642.64"

Cervical Abnormalities:

"622.5", "654.5", "654.50","654.51","654.52","654.53","654.”
